# Integrative single-cell and genetic profiling of human heart failure identifies targets for cardiomyocyte restoration

**DOI:** 10.1101/2025.10.09.25337646

**Authors:** Tore Bleckwehl, David Schumacher, Charlotte Heymanns, Sidrah Maryam, Anne-Sophie Andries, Sean J. Jurgens, Junedh Amrute, Konrad Hoeft, Xinyu Wu, Yuchen Liu, Haewon Shin, Hendrik Milting, Susanne Sattler, Kory Lavine, Michael Nyberg, Markus Bosteen, Charles Pyke, Vivek Das, Simon J. Baumgart, Rafael Kramann, Sikander Hayat

## Abstract

Heart failure encompasses a diverse group of cardiomyopathies, including myocardial infarction, hypertrophic, dilated, and arrhythmogenic forms, each defined by distinct etiologies. By integrating single-nuclei transcriptomic data from human heart tissue across these conditions, we constructed a unified atlas containing 1.8 million nuclei from 195 individuals.

The atlas reveals disease-specific cellular clusters, transcriptional changes and altered ligand-receptor interactions. We found cell states specific to the ischemic zone of myocardial infarction, explored the influence of cytokines on fibroblast cell-states, and identified etiological pathways in different cell types. The integration of summary statistics of 52 genome-wide association studies with atlas-wide gene expression highlighted genetically associated pathways involving metabolic dysregulation and ion channel dysfunction.

Implementation of an AI agent led to the identification of ZLN005, a small molecule that boosts mitochondrial biogenesis via PGC-1α, whose cardioprotective effect we validated experimentally, underscoring the utility of the atlas in early therapeutic target discovery for heart failure.

## Main

Heart failure represents a major global health burden, with ischemic heart disease as its leading cause. Dilated cardiomyopathy (DCM) accounts for the highest prevalence among non-ischemic cardiomyopathies, which exist in both idiopathic and genetic forms^1^. In contrast, hypertrophic cardiomyopathy (HCM) is the most frequently inherited cardiomyopathy. Myocardial infarction (MI) is characterized by irreversible cardiomyocyte loss and adverse ventricular remodeling, DCM by heterogeneous genetic or idiopathic defects converging on impaired contractility, fibrosis, and mitochondrial dysfunction, and HCM by sarcomeric gene mutations that induce hypercontractility^2–4^. This etiological diversity presents therapeutic challenges for each type of heart disease. For instance, regeneration and scar remodeling are promising strategies for myocardial infarction^5^, while non-obstructive therapies and fibrosis prevention have been investigated for HCM^6^. In contrast, addressing genetic variants in DCM remains difficult. Nevertheless, pathway-convergent targets may provide therapeutic benefits for both idiopathic and genetic subtypes^7^.

To better understand heart failure mechanisms, several single-nucleus RNA sequencing (snRNA-seq) studies have provided unprecedented resolution into the cellular landscape of the non-failing and failing human heart, including mapping cells across different human heart regions^8–10^, HCM^11–13^, DCM^11,14,15^, and MI^16–19^.

Currently, these conditions cannot be directly compared due to the lack of a unified framework for cell-type clustering, annotation, and downstream analyses. This hampers the identification of patient-level variation, and limits discovery of conserved and unique mechanisms in heart failure. To address this, we integrated 6 publicly available snRNA-seq datasets and standardized cell-type annotation and downstream analyses. Notably, drug targets supported by cell-type and disease-specific expression, and corroborated by genetic evidence, are associated with higher rates of clinical trial success^20^. Taking this into consideration, we further integrated the snRNA-seq data with the summary statistics from genome-wide association studies (GWAS), mapped the genetically associated genes to single-nucleus ATAC-seq and expression Quantitative Trait Locus (eQTL) and showed that this approach enables transcription-based patient stratification.

Finally, we built an AI agent that identified a small molecule ZLN005, a PGC-1α agonist, which was experimentally validated by showing its protective effects on cardiomyocytes, highlighting the activation of mitochondrial biogenesis as a convergent and therapeutically relevant pathway in heart failure. Overall, the heart atlas provides a blueprint for a more comprehensive understanding of heart failure and AI-guided target discovery.

## Results

### Construction of the adult human single-nucleus heart failure atlas

To directly compare and analyse different forms of heart failure (**Fig. 1a**), we integrated six publicly available snRNA datasets encompassing arrhythmogenic cardiomyopathy (ACM), dilated cardiomyopathy (DCM)^14,15^, hypertrophic cardiomyopathy (HCM)^11^, and myocardial infarction (MI)^16,17^, alongside non-failing heart samples (control)^8^. Datasets from adult (age >19 years) human left ventricular tissue were selected, while single-cell sequencing studies were excluded since most do not capture cardiomyocytes.

**Figure 1:**
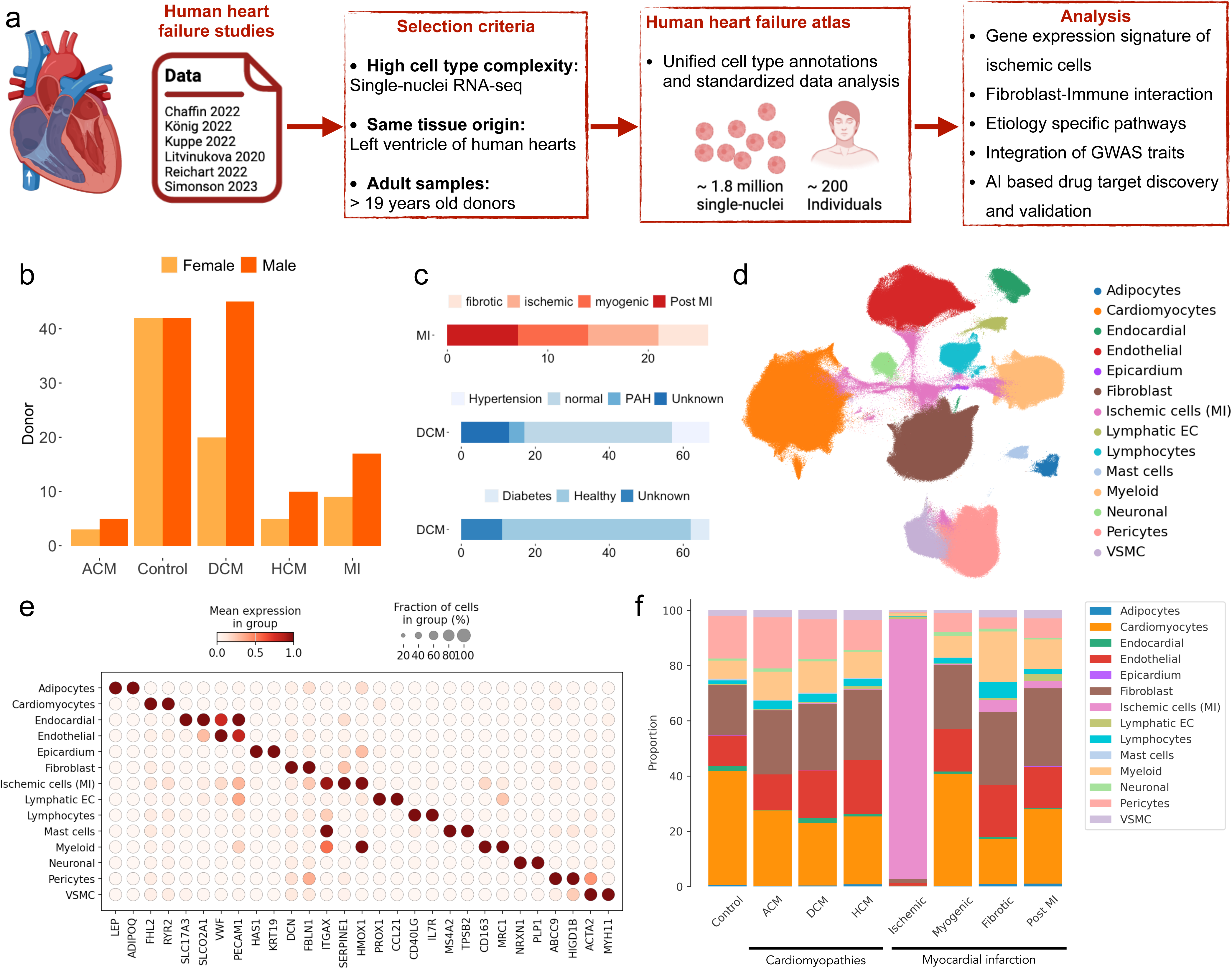
Integration of the human single-nucleus heart failure atlas. a - Overview of the integration pipeline for the human single-nuclei heart failure atlas, compiled from six independent studies. Samples derived from the adult left ventricle were filtered for high data quality and integrated using *scVI*. The resulting atlas comprises 195 patients and approximately 1.8 million nuclei, providing harmonized cell type annotations and curated metadata for cross-study comparison and downstream analysis. b - Bar plot showing the distribution of donor sex and end-stage diagnoses. Diagnoses include arrhythmogenic cardiomyopathy (ACM), dilated cardiomyopathy (DCM), hypertrophic cardiomyopathy (HCM), and myocardial infarction (MI). c - Proportional analysis of sample origin within MI subgroups (top), and prevalence of comorbidities (hypertension and diabetes) among DCM donors. d - UMAP of the integrated human heart failure atlas, colored by the major cardiac cell types identified based on marker gene expression and overlap with published annotations. e - Expression patterns of selected marker genes across major cardiac cell types in the atlas. f - Cell type compositional changes inferred by *scCODA*, comparing donor controls, cardiomyopathies, and different myocardial infarction sample sites and stages.

After a rigorous quality filtering process including doublet removal, the atlas contained 1,751,865 nuclei of human left ventricle tissue from 195 individuals with a median of 8,250 nuclei per donor (**Supplementary Fig. 1a**), representing 42 female and 42 male non-heart failure donors as controls, and 36 female and 75 male specimens with various heart failure etiologies (**Fig. 1b**). Clinical meta data from the individual studies were harmonized, conserving the original sample sites of the myocardial infarction samples, comorbidities, body mass index (BMI) and others (**Fig. 1c, Supplementary Fig. 1b, Supplementary table 1**). As for other single-cell atlas^21,22^, scVI^23^ was applied for batch correction, balancing sample effects from different studies (**Supplementary Fig. 1c**). Batch-effect corrected datasets were clustered and annotated based on cell-type specific marker gene expression, revealing major cardiac cell types at their expected abundance. Our cell type annotations provide a unified description that overlapped with the original annotations from the different integrated studies (**Fig. 1d-e, Supplementary Fig. 1c-d, Supplementary table 2**).

Notably, rare cell types, such as adipocytes, lymphatic endothelial cells and neuronal cells were represented together with major cardiac cell types. Beyond known cell populations, the atlas revealed an ischemic cell cluster, which was transcriptionally different from all other main clusters and enriched in samples from the ischemic zone (**Fig. 1f**). Besides, compositional analysis revealed that cardiomyocytes showed a reduced proportion in heart failure in general as well as in individual heart tissues from DCM patients (**Fig. 1f, Supplementary Fig. 1f**).

### Gene expression signature and pathways of acute human ischemia

The main cell type clustering highlighted transcriptional differences in ischemic cells sampled from the ischemic zone of acute MI (**Fig. 1d**). Differential expression analysis of the ischemic cell cluster, revealed enriched markers (Plasminogen activator inhibitor-1: *PAI-1/SERPINE1*, Stabilin1: *CLEVER-1/STAB1*). Unlike the markers of other major clusters, ischemic cell markers frequently displayed expression across multiple cell types (**Fig. 2a**). Subclustering of the ischemic cluster revealed nine subclusters representing ischemic counterparts of the major cardiac cell types, including ischemic cardiomyocytes, mononuclear phagocytes and fibroblasts with increased *COL3A1* and *COL1A2* expression and reduced gene expression of fibroblast markers such as decorin (*DCN*) (**Fig. 2b, Supplementary Fig. 2a**).

**Figure 2:**
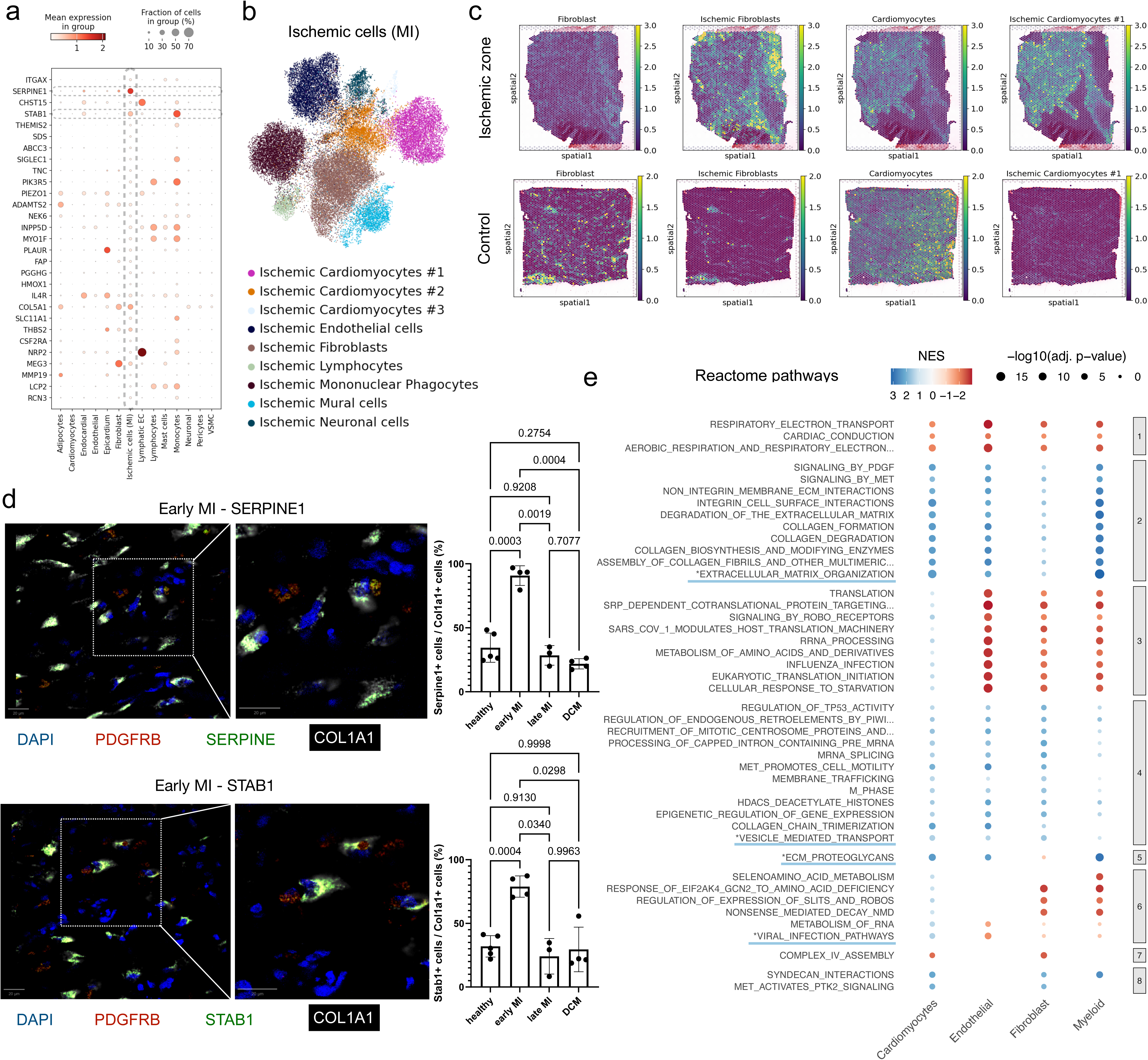
Gene signature of the ischemic cell cluster. a - Marker gene expression of the ischemic cell cluster vs. all other nuclei. Experimentally validated transcriptional markers *STAB1* and *SERPINE1* are highlighted. b - Subclustering of the ischemic cell cluster showing distinct cell types in the ischemic zone of patients with acute myocardial infarction. c - Cell state mapping of ischemic cardiomyocytes and fibroblast states to spatial transcriptomics slides of 10X Genomics Visium samples sampled from the ischemic zone and from control donor heart tissue. d - Examples of the RNA-scope experiments for the presence of *STAB1* and *SERPINE1* mRNA in early myocardial infarction, including a zoom-in for the co-localization with *COL1A1*. The summary for the quantification of co-occurances of *COL1A1* and *STAB1* or COL1A1 and *SERPINE1* are shown for the indicated heart failure conditions. Dots represent individual patients, the barplot the mean for each condition and p-values were calculated from one-way ANOVA (equal variances) or Welch ANOVA (unequal variances). e - Reactome gene-set enrichment analysis from the pseudobulk differential expression analysis of indicated ischemic clusters versus the corresponding cluster from control donors. The cardiomyocytes of the ischemic cell clusters were considered as one ischemic cardiomyocyte cluster.

For orthogonal validation of the ischemic cell state, we mapped the gene signatures from the ischemic and non-failing control cluster to spatial transcriptomics data of different sites of heart tissue samples from patients with acute MI^16^. This showed that the abundance of non-failing major cardiac clusters is reduced in the ischemic zone, while different ischemic clusters showed increased abundance within the ischemic zone of acute infarction, indicating that the defined ischemic cell clusters correspond to the histopathologically defined distinct ischemic regions (**Fig. 2c, Supplementary Fig. 2b**). Additionally, to mitigate inference from ambient RNA, raw gene expression counts were compared with cellbender corrected counts for predicted ambient RNA contaminations^24^, showing only negligible changes in the gene expression signature of ischemic cells (**Supplementary Fig. 2c**). While markers such as *SERPINE1* were ubiquitously upregulated, other markers indicated a cell type specific ischemic response, e.g. *SLC11A1* in mononuclear phagocytes and *MEG3* in ischemic fibroblasts. (**Supplementary Fig. 2d**).

Finally, the ischemic gene signature was experimentally validated by RNA-scope experiments for *SERPINE1* and *STAB1,* confirming the presence of both transcripts in early human MI and their co-localization with *COL1A1* compared to non-failing controls as well as late MI and DCM (**Fig. 2d, Supplementary Fig. 2e**). Furthermore, the expression changes of the top differentially expressed genes in human MI (*e.g. ADAMTS2*) were confirmed by scRNA-seq from isolated PDGFRβ positive fibroblasts of mice with chronic MI by permanent ligation of the left anterior descending artery (LAD) (**Supplementary Fig. 2f**).

Comparing gene expression of ischemic cells against non-failing controls revealed downregulation of non-coding RNAs in cardiomyocytes and several pathway alterations (**Fig. 2e, Supplementary Fig. 2g, Supplementary table 3**). In particular, the activity of the respiratory electron transport pathway was decreased across all ischemic cell types, while pathways including extracellular matrix organization and ECM proteoglycans, involving SERPINE1, and vesicle-mediated transport and viral infection, involving STAB1, were upregulated.

### Patient profiling revealing different cell state compositions

To better resolve cellular and patient heterogeneity, we performed subclustering of the major cardiac cell types and generated patient representations by aggregating pseudobulk RNA expression profiles which revealed six patient clusters that could be further categorized into three patient clusters of non-failing controls and three patient clusters with different forms of heart failure derived from independent studies (**Fig. 3a, Supplementary Fig. 3a**–b).

**Figure 3:**
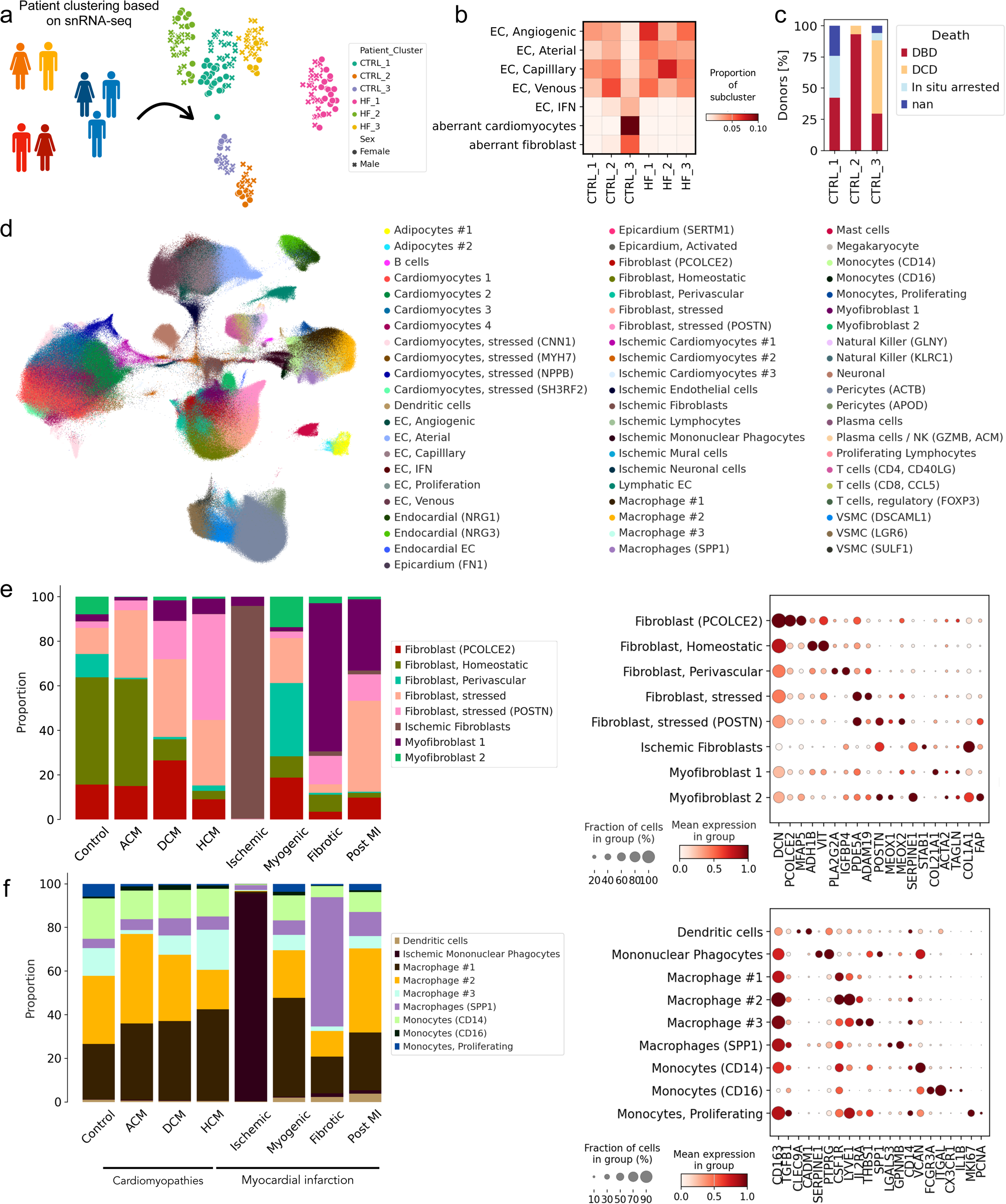
Patient profiling, subclustering and cardiac cell type heterogeneity. a - Overview of patient clustering based on aggregated single-nucleus RNA-seq expression profiles. Patients were grouped via Leiden clustering, colored by cluster identity and donor sex is indicated by shape. b - Matrix plot showing the subcluster of endothelial cells as well as aberrant fibroblasts and aberrant cardiomyocytes as the mean proportion in each patient cluster. c - Percental distribution of disease diagnosis across patient clusters (left). For clusters containing control samples, the cause of death is shown on the right. DBD: donation after brain death; DCD: donation after cardiac death; nan: unknown. d - UMAP visualization of subclustering for major cardiac cell types. e - *scCODA*-based compositional analysis of fibroblast cell subcluster across conditions, including non-failing controls, cardiomyopathies, and myocardial infarction at different stages (left). Gene expression of selected markers for each fibroblast subcluster (right). f - Cell type compositional changes inferred by *scCODA* for myeloid subclusters across conditions, including non-failing controls, cardiomyopathies, and myocardial infarction at different stages (left). Gene expression of selected markers for each myeloid subcluster (right).

Subclustering of the major cell types, revealed populations with compromised cell-type marker expression and high ribosomal/mitochondrial gene content, which were labeled as aberrant cardiomyocytes and fibroblasts. These aberrant clusters were exclusively enriched in one patient cluster (CTRL_3), while the other patient clusters varied in the cell type proportions with one patient cluster showing higher proportions of myeloid cells (**Fig. 3b, Supplementary Fig. 3c**). We further noticed that samples in CTRL_3 patient cluster consists of donors that were collected post cardiovascular death (DCD), suggesting these populations may represent technical artifacts (**Fig. 3c**). Given the difficulty of obtaining reliable non-failing control samples, we excluded both aberrant clusters and donors from CTRL_3 from further analysis (excluding 7 female and 11 male donors).

### Subclustering for assessment of subtype heterogeneity

Subsequent subclustering of the cleaned dataset identified 64 subclusters with distinct compositions across different types of heart failure (**Fig. 3d).** Within the cardiomyocytes, cardiomyocytes 1 (expressing *CORIN*) represented the dominant population in non-failing controls, whereas stressed cardiomyocyte subclusters were markedly enriched in specific donors. Cardiomyocytes 4 (expressing *ADRA1A*) were selectively enriched in a small patient subgroup, while cardiomyocytes 2 and 3 (with increased MT- or *PLCE1* gene expression, respectively) were broadly present among cardiomyopathies (**Supplementary Fig. 3d-f**). Similarly, fibroblasts showed a homeostatic fibroblast cluster that showed a high proportion in non-falling controls and gene expression of classic fibroblast markers such as *DCN*, whereas stressed fibroblasts (expressing *PDE5A*, *ADAM19*) and *PCOLCE2* positive fibroblasts were enriched in patients with DCM (**Fig. 3e**). For the DCM-related fibroblasts subtypes we noted a patient-level continuum in DCM patients (**Supplementary Fig. 3g**). Interestingly, *POSTN* positive stressed fibroblasts were found in high proportions in patients with HCM, while myofibroblast1 (expressing *ACTA2*, *TAGLN*) were enriched in fibrotic samples of acute MI or post MI patients. Perivascular fibroblasts, identified by the expression of *PLA2G2A*^18^, were abundant in a subset of patients, potentially reflecting site-specific alterations in the sampled tissue.

Overall, there was a high overlap with the subclusters of individual studies for cardiomyocyte and fibroblast sub types, which illustrates our harmonization of diverse fibroblast populations into a unified and comparable annotation (**Supplementary Fig. 3f-g**). For example, the *PCOLCE2* positive fibroblasts mainly originated from the vFB4^14^ and FB-ZBTB7C^11^ clusters in their respective authors’ annotations.

Additionally, condition specific patterns were observed: natural killer cells (*GNLY*) were increased in DCM, angiogenic endothelial cells were enriched post myocardial infarction (post MI) and NRG1-positive endocardial subclusters predominated in DCM (**Supplementary Fig. 4a, Supplementary table 4**). The presence of specific *NRG1*- and *NRG3*-expressing endocardial cell populations is noteworthy, as neuregulins are known to play crucial roles in cardiomyocyte survival, proliferation, and cardiac function^25^. Within myeloid cells, macrophage clusters 1-3 exhibited altered expression of the general macrophage marker *CSF1R*, the resident macrophage marker *LYVE1* and the pro-fibrotic marker *THBS1* with the latter specifically expressed in macrophage 3, a subtype with high proportion in HCM (**Fig. 3f**). Additionally, monocytes and dendritic cell clusters were present across all conditions, and the proportion of *SPP1* expressing macrophages was elevated in fibrotic MI samples.

Furthermore, differential expression analysis revealed that etiological differences were reflected in gene expression changes (**Supplementary table 5-6**), providing opportunities to investigate heart failure-specific mechanisms and to identify common or condition-specific therapeutic targets.

### Dysregulation in cell-cell communication

To explore ligand-receptor interactions, we applied MultiNicheNet^26^ and CrosstalkR^27^. CrosstalkR showed altered cell-cell communication in different heart diseases with dysregulated interactions between cardiomyocytes and vascular smooth muscle cells (VSMC) in HCM and DCM, while in DCM interactions of cardiomyocytes with neuronal cells, fibroblasts and endothelial cells were additionally increased. In acute MI, putative cell-cell communication between cardiomyocytes, fibroblasts, VSMC and pericytes was enriched (**Fig. 4a, Supplementary Fig. 5a, Supplementary table 7**). To elucidate shifts in ligand-receptor interactions in different heart diseases and extract etiology-specific interactions, we clustered the cell-cell interactions by their disease enrichment score to highlight universal and disease-specific ligand-receptor interactions (**Fig. 4b**). We identified putative communication of COL1A1/2 in ischemic conditions, NRG1 to ERBB4 signaling in ischemic conditions and DCM, NCAM1-CACNA1C interactions in DCM and CALM2-PDE1C interaction in the HCM **(Supplementary Fig. 5b**). Etiologic-specific interactions were predicted for SERPINE1 and PDGFRA/B in ischemic conditions, COL6A6 in DCM as well as PTN-interactions in HCM and DCM (**Fig. 4b**).

**Figure 4:**
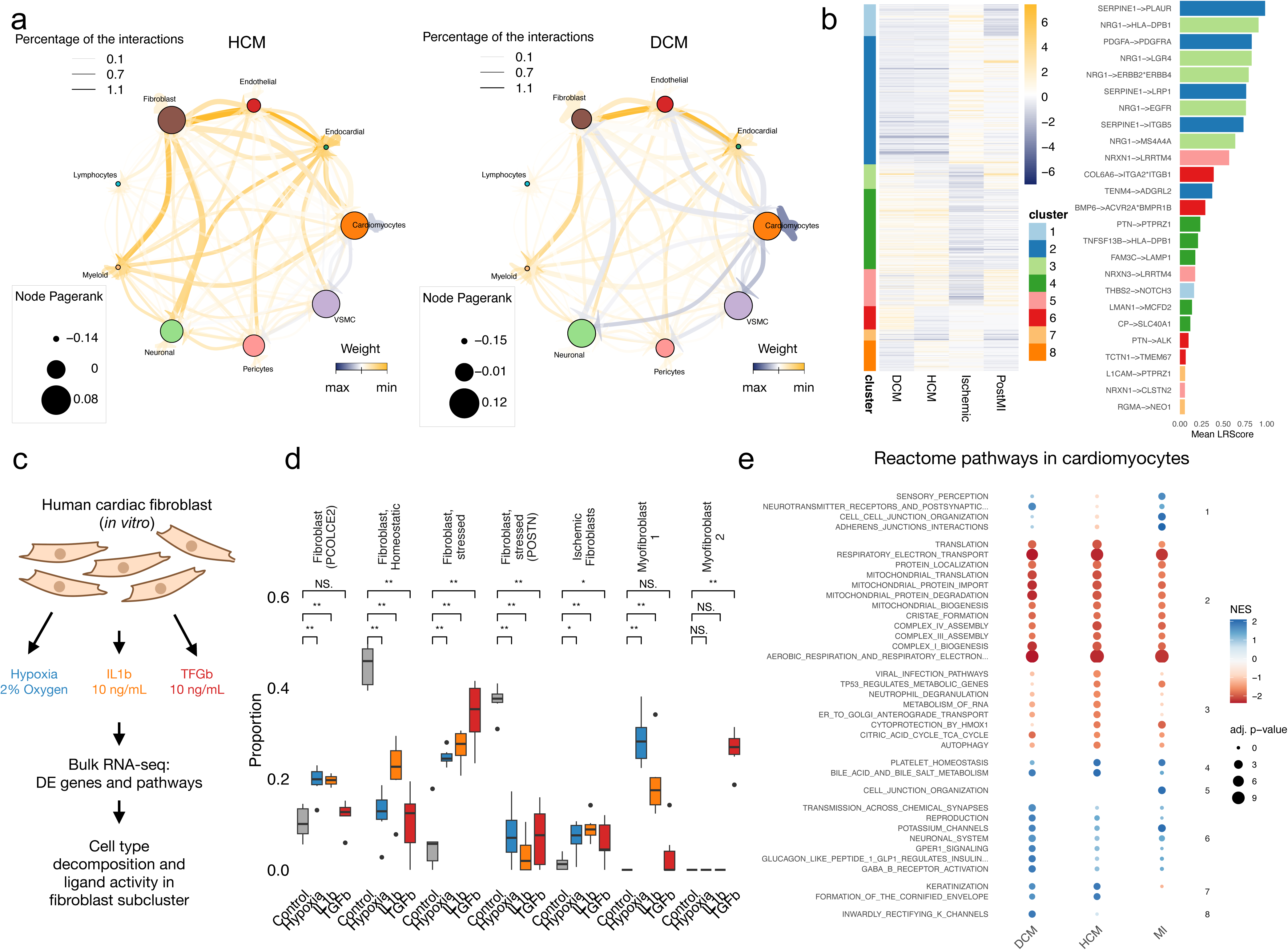
Divergent cell-cell communication across heart failures and insights into immune cell cross talk with different fibroblast states. a - CrosstalkR results for the cell-cell-communication in the different heart failure conditions. The weighted interactions for HCM on the left and DCM on the right. For the analysis the consensus interactome of liana was extended by the GPCR module. b - Clustering of the z-scored LRScore for each condition of the different heart failure diseases on the left. Unique ligand-receptor interactions for each cluster are shown on the right with the LRScore summarized over all cell types. c - Schematic overview of the experimental design for the in vitro treatment of cardiac fibroblast with IL-1β, TGF-β and hypoxia to study the gene expression changes by bulk transcriptomics. d - Cell type decomposition with *Bisque* for each of the in vitro fibroblast treatments based on the fibroblast subcluster gene signature of the human heart atlas. e - Analysis of cardiomyocyte enriched reactome pathways and their changes in dilative and hypertrophic cardiomyopathy as well as myocardial infarction (MI). The pathways are clustered by similarity of the normalized enrichment score (NES).

MultiNicheNet showed prioritized ligand activity influenced by multiple cell types including endothelial cells in post MI and fibroblasts in DCM (**Supplementary Fig. 5c**). Potential endothelial cell interaction in post MI pointed towards angiogenesis and vascular repair (VEGFC, FGFR1). Predicted fibroblast interactions in DCM were more widespread, involving collagen interaction, TGF-β signaling and interaction of IL16 with KCNJ4 in cardiomyocytes as well as NPPB-NPR3 interactions between cardiomyocytes.

### Cytokine stimulation of fibroblasts and ion channel dysregulation in cardiomyocytes

To understand the influence of different cytokines on fibroblast subtypes, human cardiac fibroblasts were exposed to IL-1β, TGF-β and hypoxia followed by bulkRNA-seq to measure gene expression changes (**Fig. 4c**). Each stressor showed the expected pathway dysregulation, namely signaling by interleukins for IL-1β, signaling by TGF-β family members for TGF-β stimulation, and cellular response to hypoxia under hypoxic conditions (**Supplementary Fig. 5d-e, Supplementary table 8**). To test whether fibroblast subtype expression profiles were mimicked *in vitro*, we applied a regression-based deconvolution approach^28^ and estimated the fibroblast subtype proportions per condition (**Fig. 4d**). Untreated samples were enriched for homeostatic fibroblasts, while all stimulations increased ischemic and myofibroblast signatures. TGF-β showed the highest proportion of DCM-related stressed fibroblast signal (**Fig. 3f**), but had a weaker effect on DCM-related *PCOLCE2*-positive fibroblast or stressed fibroblast (*POSTN*) linked to HCM.

Next, we explored cell-type and etiology-specific pathways in major cardiac cell types and heart failure. Cardiomyocyte-specific pathways (**Fig. 4e, Supplementary table 9**) showed eight different categories related to increased cell-cell communication activity in DCM and MI, reduced mitochondrial biogenesis and respiratory electron transport in all heart failure conditions, downregulation of autophagy in HCM as well as upregulation of potassium channels, neurotransmitter receptor and postsynaptic signal transmission (*e.g. PRKCA, CREB1*), and GLP1 regulated insulin secretion in DCM. Bulk RNA-seq of *in vitro* cultured cardiomyocytes exposed to doxorubicin showed upregulation of potassium channels and therefore indicating a stress-induced remodeling (**Supplementary Fig. 5f**). Notably, potassium channels such as *KCNJ4*, displayed an overall increase in the percentage of expressing cells across the DCM patients. In contrast, the proportion of GLP1R-expressing cardiomyocytes was heterogeneous across individuals, with a subset of patients showing higher levels in DCM (**Supplementary Fig. 5g**). Because GLP1R expression is typically enriched in the sinoatrial node (SAN)^29^, we compared the expression of potassium channels, GLP1R pathways and established pacemaker-associated markers (*e.g. HCN4*) across the conditions of the atlas, published cardiac niches subtypes^9^, and doxorubicin-treated cardiomyocytes (**Supplementary Fig. 5h)**. This confirmed the absence of pacemaker markers in the ventricular cardiomyocytes and suggests a stress-induced expression dysregulation of potassium channels in the DCM patients.

### GWAS integration with snRNA-seq reveals genetically associated pathways

Cardiomyopathies arise from a complex interplay of genetic predisposition, idiopathic factors, and comorbid conditions. Genome-wide association studies (GWAS) have identified common variants but a comprehensive causal understanding of the affected pathways and their cell-type specificity remains limited. To overcome this, we applied the polygenic regression framework of scPagwas^30^ to integrate GWAS summary statistics with snRNA-seq data. A polygenic regression model for the beta variance from GWAS and the pathway activity from snRNA-seq was fitted, yielding a genetically associated pathway score and thus enabling a polygenic association. We systematically evaluated polygenic contributions in cardiomyopathies by analyzing summary statistics from 52 GWAS traits derived from ∼40 million cases in the EMBL-EBI GWAS catalog^31^ (**Fig. 5a, Supplementary Fig. 6a, Supplementary table 10**). Prioritized genes for a trait were determined by ranking the Pearson correlation coefficients (PCC) from the correlation of genetically associated pathway activities and the gene expression of a given gene across all individual cells, highlighting genes like inositol 1,4,5-trisphosphate receptor (*InsP3R2, ITPR2*)^32^, splicing factor Quaking (*QKI*)^33^ and others that have been previously genetically associated with DCM or HCM (**Fig. 5b, Supplementary Fig. 6b, Supplementary table 11**).

**Figure 5:**
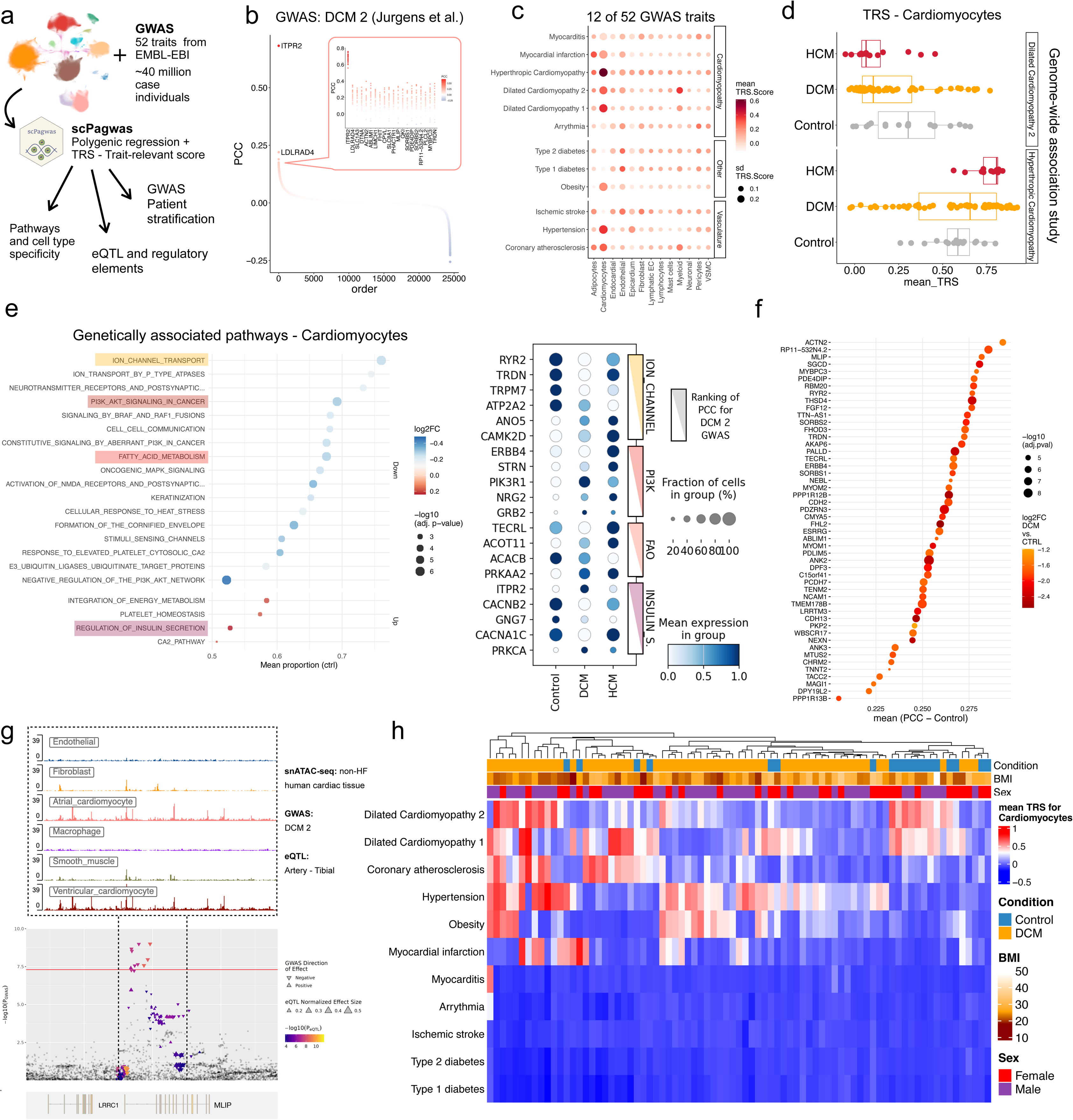
GWAS association reveals polygenic expression changes in different cell types and pathways for primary cardiomyopathies. a - Overview of the genetic linkage to the human heart failure atlas, applied to 52 genome-wide association studies (GWAS). The trait relevant scores (TRS) were calculated with scPagwas and downstream analysis were performed for changes in genetically associated pathway activities, regulatory analysis of GWAS associated genes and patient stratification. b - GWAS DCM 2 relevant genes ranked by the Pearson correlation coefficient (PCC) as mean from patients with non-failing hearts. The top ranked genes are highlighted in the box, where each dot denotes the ranking and PCC from non-HF individuals. c - Trait-relevant score for 12 selected traits. The TRS-scores were calculated per patient with scPagwas and aggregated by the mean value per cell type cluster from the patient of the control as well as dilative and hypertrophic cardiomyopathy. The size denotes the standard deviation of the mean of all patients. d - Comparison of the trait-relevant score (TRS) in cardiomyocytes for different conditions and the indicated GWAS. Each dot represents the mean TRS from all cardiomyocytes of an individual. e - Trait-relevant pathways for DCM and cardiomyocytes arranged by the mean proportion of trait-associated pathways (pathway-level coefficient β > 0) from the controls and colored by the log2 fold changes of DCM vs non-HF for the mean proportion of trait-associated pathways from all individuals. For selected pathways the top genetically associated genes for that pathway and their mean expression within the indicated condition from the snRNA-sea heart failure atlas is shown. f - Pearson Correlation Coefficient (PCC) for genes associated with both DCM 1 and DCM 2 GWAS and the log2FC in the PCC for DCM 2 vs control. g - Manhattan plot for −/+ 200kb of the *MLIP* gene for the GWAS for DCM 2. Overlapping SNPs with the expression quantitative trait locus (eQTL) from the tibial artery are highlighted by the effect size. For the dashed region with significant SNP, the single-nuclei ATAC-seq data from Hocker *et al.* 2021 is shown for the indicated cell types. h - Heatmap of the mean trait-related score per patient (x-axis) in cardiomyocytes for different traits (y-axis). The data was clustered by columns and rows and clinical meta data for the condition, sex and body-mass-index (BMI) is colored on top of the heatmap.

The top-ranked genes of the PCC were combined into a gene set, which by cell-scoring against random genes is represented as a single value, defined as trait-relevant score (TRS). Using this approach, we summarized the TRS from patients including non-failing donors as control and patients with HCM or DCM, which showed that cardiomyocytes had the highest TRS for DCM and HCM GWAS traits, respectively (**Fig. 5c, Supplementary Fig. 6a**). Notably, there was an increased TRS for the GWAS traits of obesity and hypertension in cardiomyocytes as well. For cardiomyocytes, the TRS for the DCM GWAS was lower in the cardiomyocytes from atlas patients with DCM/HCM, indicating that genes that are genetically associated with DCM mostly were downregulated in cardiomyocytes of DCM patients (**Fig. 5d**). There was no clear indication for differences in the patient genotypes (**Supplementary Fig. 6c**). Next, we focused on the genetically associated pathways estimated with scPagwas and calculated the proportion of cells with positive associations for each pathway. Since the TRS indicated a strong effect in cardiomyocytes, we prioritized pathways with high cardiomyocyte specificity, quantified by normalized enrichment across cell types, and showed that this approach highlights cardiomyocyte-specific pathways for DCM and HCM (**Supplementary Fig. 6d-e**). Cardiomyocyte specific genetically associated pathways in DCM involved ion channel transport, with calcium transporters (*RYR2, TRPM7, ATP2A2*) achieving the highest rank for their genetic pathway association (**Fig. 5e**). Other genetically associated pathways in cardiomyocytes for DCM involved PI3K-AKT signaling with ERBB4 whose interaction to NRG1 was predicted to be DCM related (**Fig. 4b**), fatty acid metabolism involving TECRL and regulation of insulin secretion, showing genetic association and downregulation in DCM for other calcium transporters (*CACNA1C*).

Independent from the pathways, we analysed genes ranked by PCC for HCM and DCM GWAS traits, revealing known candidates like *FHOD3* for DCM and *MYBPC3* for HCM (**Fig. 5f, Supplementary Fig. 6f**). The top-ranked genes also included Muscular LMNA interacting protein (*MLIP*)^34^ and other sarcomere and cytoskeletal genes such as *ACTN2*, *PDE4DIP* and *FHOD3* with known implication in DCM and significant loci in GWAS^35^. To validate top-ranked genes, we mapped expression Quantitative Trait Locus (eQTL) from Genotype-Tisse expression (GTEx) biobank and single-nuclei chromatin accessible data from non-failing human heart tissue^10^ to the SNPs and related loci of DCM 2 GWAS gene. Indeed, a significant proportion of GWAS SNPs for *MLIP* and *FHOD3* overlapped with eQTL of a negative effect size, indicating a negative impact on the gene expression and mapping with chromatin accessible sites that indicate *cis*-regulatory elements in the intronic regions (**Fig. 5g, Supplementary Fig. 6g-h**).

Lastly, to obtain further insights into comorbidities from GWAS, we summarized the TRS score for cardiomyocytes of selected GWAS traits in non-failing controls and DCM patients from the heart atlas. Clustering revealed a potential approach to transcriptionally stratify the patients as DCM patients differed in their interference with TRS from other GWAS such as hypertension and obesity that correlated well with the clinical meta data of the patients (**Fig. 5h, Supplementary Fig. 6i**).

Overall, we show that the integration of snRNA-seq and summary statistics from GWAS can reveal genetically linked pathways and enable transcriptional patient stratification.

### AI agent for drug target discovery of ZLN005 and its experimental validations

Our analyses from differential gene expression (**Fig. 3**), cell-cell-communication, pathway analysis, *in vitro* cultured cardiomyocytes (**Fig. 4**) and integrative genetics analyses (**Fig. 5**) pointed out different layers of changes in the potassium and calcium handling. To elucidate it further, we extracted the subcellular location of calcium channels orthogonally identified in our analyses and selected top genes with the highest TRS score for DCM. It showed that the calcium channels with a high TRS were highly expressed in cardiomyocytes and localized preferentially in the sarcolemma and sarcoplasmatic reticulum (SR), while it was less evident for synaptic or mitochondrial calcium channels (**Fig. 6a,b**).

**Figure 6:**
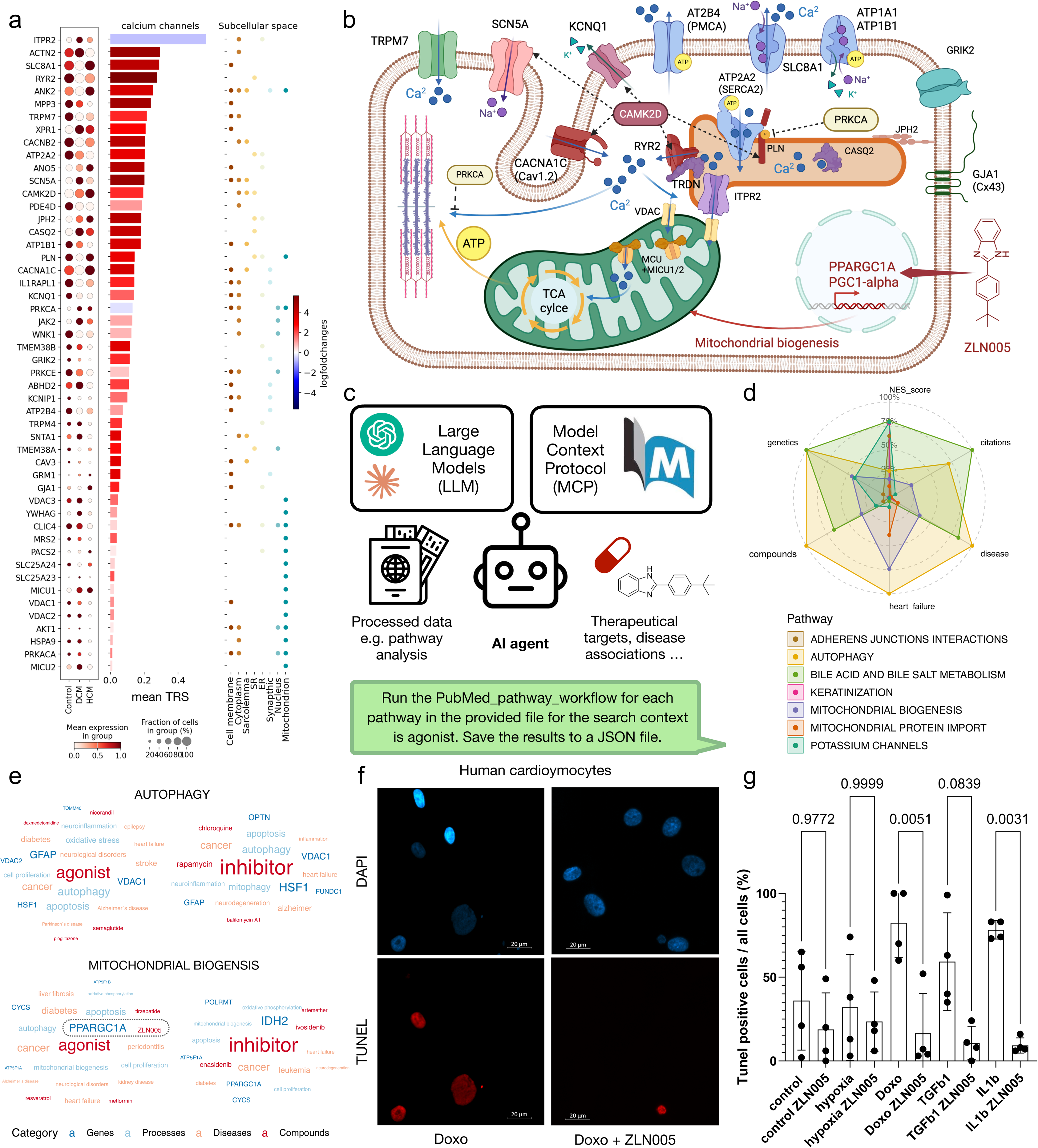
AI agent for customized drug target discovery and its experimental validations. a - Overview of the top uniprot calcium channels (and associated proteins) by subcellular locations. Expression of the calcium channels in the human heart failure atlas shown on the left, the genetic association expressed by the mean TRS score for each gene on the right. The coloring indicates the specificity for cardiomyocytes defined by the log2foldchange of the gene expression in cardiomyocytes vs. all other cells. On the right are indicated the subcellular locations from the uniprot database for each calcium channel. b - Schematic illustration of the calcium channels, their subcellular localizations and calcium flux within a cardiomyocyte. The small molecule 2-(4-tert-butylphenyl)-1H-benzimidazole (ZLN005) activates the transcription of peroxisome proliferator-activated receptor gamma coactivator 1-alpha (PGC-1α, PPARGC1A) promoting the biogenesis of mitochondria. c - Overview of the AI agentic approach in which a large language model coordinates a pubmed search with a model context protocol (MCP) server. For pre-selected pathways, an OR query across all member genes retrieves relevant literature, and the agent extracts diseases, therapeutic compounds, and biological processes to generate pathway-level summaries. d - Radar plot showing the analysis of the AI agent output from the pubmed search for cardiomyocyte specific pathways in heart failure. The occurrences of general diseases, heart failure, compounds and genetic related studies as well as the overall found studies (citations) are min-max normalized, while the NES score was scaled from −3 (equal to 0%) to +3 (equal to 100 %). e - Word frequency plots derived from the AI agent PubMed search of the cardiomyocyte specific pathways for autophagy and mitochondrial biogenesis in the context search of agonist (left) and inhibitor (right) were colored by the different categories. Highlighted are the most frequently occurring gene name PPARGC1A (PGC-1α) and the small molecule ZLN005 from the agonist search with the genes of the mitochondrial biogenesis pathway. f - Representative DAPI (top) and TUNEL staining (bottom) of untreated cardiomyocytes (control), cardiomyocytes treated with doxorubicin and doxorubicin and the PGC-1α agonist ZLN005. g - Summary of the experimental validation for ZLN005 and different treatments of *in vitro* human cardiomyocytes, including untreated cardiomyocytes as a control. On top are displayed the p-values for the treatment and the recovery with ZLN005 from a one-way ANOVA for multiple comparisons.

To assess the translational value of our atlas, we implemented an agentic workflow in which a large language model (LLM) orchestrates pathway-specific PubMed searches via a Model Context Protocol (MCP) server. This provides a standardized, auditable interface that makes LLM-driven literature retrieval reproducible and transparent (**Fig. 6c**). The agent thereby interprets the current literature of the genes involved in the pathway for a defined research context to identify relevant compounds and drug targets. To test our agent, we applied it to the cardiomyocyte specific pathways (**Fig. 4e**) in the context of existing agonists and inhibitors. Subsequent analysis of the agent response revealed a high frequency of heart failure related publications related to mitochondrial biogenesis and autophagy pathways (**Fig. 6d, Supplementary Fig. 7a**). For the mitochondrial biogenesis pathway, IDH2 dominated in inhibitor-linked search and PPARGC1A in agonist-linked search (**Fig. 6e**). For the genetically associated pathways (**Fig. 5e**), the AI agent surfaced multiple studies with heart failure and therapeutics such as the ERBB4 agonist JK07 (**Supplementary Fig. 7b**,c). We reasoned that a general drug target could support cardiac reinstating, addressing coherent improvement, and thus focused the experimental validation of an agonist for a downregulated pathway. Given the high energy demand and coupling of mitochondrial output to calcium handling, we focused on the gene (PPARGC1A) and compound (ZLN005) with the highest occurrence. ZLN005 has been reported to enhance PPARGC1A-dependent transcription programs essential for mitochondrial biogenesis^36^. Notably, ZLN005 is not yet linked to target PPARGC1A in public databases for bioactive molecules (e.g. PubChem, ChEMBL), indicating the feasibility to capture emerging compounds with the AI agent since ZLN005’s effect has been studied in liver^37^, kidney^38^ and cerebral infarction^39^.

Expression profiling of the genes from the mitochondrial biogenesis pathway showed elevated expression in cardiomyocytes, consistent with their energetic requirements for contraction (**Supplementary Fig. 7d**)^40^. In agreement with the pathway analysis, reduced expression was observed in cardiomyocytes across different heart failure etiologies (**Supplementary Fig. 7e**), aligning with a metabolic shift from oxidative phosphorylation to glycolysis, which was confirmed in the border zone of human MI by spatial transcriptomics (**Supplementary Fig. 7f**).

Next, we evaluated the compound ZLN005 for its ability to preserve cardiomyocytes when exposed to damaging stimuli. For this purpose, human cardiomyocytes were cultured under hypoxia or exposed to IL-1β, TGF-β or doxorubicin. All exposures showed elevated apoptotic cardiomyocytes with increased percentage of TdT-mediated dUTP-biotin nick end labeling (TUNEL) (**Fig. 6f, Supplementary Fig. 7g**). Importantly, the addition of the PPARGC1A agonist ZLN005 could significantly reduce cell death to levels observed in untreated cardiomyocytes, indicating the protective effect of functional mitochondria (**Fig. 6g**). Additionally, the expression of known cardiac injury markers *ANKRD1* and *NPPB* showed increased expression under all conditions (**Supplementary Fig. 7h**, i).

In summary, we localized GWAS-informed factors of calcium flux in cardiomyopathies, demonstrated a lightweight LLM-powered AI agent that identified relevant compounds to tackle pathways of our heart atlas.

## Discussion

Here, we provide an integrated human heart failure atlas of publicly available single-nuclei RNA-seq data. Our unified cell type clustering, annotation and meta data harmonization enable comparative analyses across studies. We found that nuclei sampled from the ischemic zone displayed a distinct gene signature across all cell types, showing increased expression of *SERPINE1* and *STAB1*. SERPINE1 (PAI-1) is induced by hypoxia and modulates the TGF-β axis, thereby promoting matrix accumulation and fibrosis/tissue remodeling after injury^41^ and *STAB1* expression is an early marker of macrophages in MI^42^. Interestingly, the fibrotic region of MI, was dominated by *SPP1*-positive macrophages linked to MI^43^ while other divergent immune cell subtypes impact fibroblast cell states by macrophage-fibroblast cross-talk^44,45^. Importantly, we observed that myofibroblasts were highly present in MI, *PDE5A*-positive stressed fibroblasts and *PCOLCE2*-positive fibroblasts were enriched in patients with DCM and stressed fibroblasts (*POSTN*) in HCM. *in vitro* culture of fibroblasts exposed to TGF-β, IL-β and hypoxia stimulated fibroblast differentiation resembling the myofibroblasts and DCM-related stressed fibroblasts^46^. However, the DCM-related *PCOLCE*-positive fibroblasts that appeared on a spectral ratio to the DCM-related *PDE5A*-positive fibroblasts in individuals, and HCM-related stressed fibroblasts were less represented by these modulators, which might indicate an independent stimulus for this fibroblast subtypes. Besides, NRG1-positive endocardial subclusters were predominated in DCM which might indicate supportive effects, considering the binding of NRG1 to ERBB4 on cardiomyocytes, engaging PI3K-AKT signaling and suppression of apoptosis^47^. Consistently, we predicted the NRG1-ERBB4 interaction in the cell-cell-communication analysis for DCM, together with putative interaction of interleukins (IL16), being expressed in fibroblasts, and potassium channels (KCNJ4) of cardiomyocytes as well as adhesion signaling (NCAM1) and calcium channels (CACNA1C) in cardiomyocytes. Thereby, neuronal interleukin modulation of multiple potassium channels^48^, might hint towards analogous mechanisms in injured cardiomyocytes.

The pathway analysis in cardiomyocytes pointed to two alterations with potential relevance to excitability. Firstly, GLP1R transcripts were present in a subset of ventricular cardiomyocytes, as previously noted at the mRNA level^49^, although GLP-1R is classically enriched in the SAN^29^. In a recent clinical trial^50^, treatment with the GLP-1R agonist semaglutide was found to reduce the risk of cardiovascular events. The mechanisms underlying this protective effect are likely to be linked to reductions in body weight, waist circumference, HbA1c, and hsCRP^51^, with the current findings of a low expression of GLP-1R in ventricular cardiomyocytes indicating that signalling via these receptors are of limited relevance. However, the samples of the heart atlas are end-stage heart failure and the direct impact on cardiomyocytes might warrant further research in especially patients with advanced cardiac disease, which then might help stratify treatment response to GLP-1R agonists.

Secondly, we found a broad upregulation of potassium-channel pathway genes across DCM patients, which aligns with repolarization changes in failing ventricles^52^. Its *in vitro* recapitulation after doxorubicin^53^ exposure, implicates cardiac injury pathways, stressing cardiomyocytes and disrupting potassium currents.

Importantly, we modeled pathway-level polygenic GWAS^30^ scores, highlighting a coordinated burden on calcium handling pathways, implicated PI3K-AKT signaling, including *ERBB4*, and elements of fatty-acid metabolism, including trans-2, 3-enoyl-CoA reductase-like (*TECRL*), which has been linked to mitochondrial respiration in a PI3K-AKT-manner^54^. Polygenic associations were shown for multiple genes encoding core determinants of the cardiac action potential (**Fig. 6b**), which except for SCN5A (depolarization phase), were transcriptionally downregulated. The downregulated genes included calcium channels (CACNA1C, RYR2) of the plateau phase and potassium channels (KCND3, KCNQ1) of the depolarization phases, the re-uptake of calcium into the SR (ATP2A2/SERCA2 regulated by PLN) and diastolic ion-balancing membrane channels AT2B4, SLC8A1 and ATP1B1.

This coordinated downregulation likely represents an adaptive attempt to dampen excitation-contraction coupling and limit injurious ion fluxes under stress^55^, which is consistent with the upregulation of CASQ2 (regulating calcium release from SR) and MICU1/MICU2^56^.

Interestingly, PRKCA and CAMK2D were upregulated. PRKCA is a negative modulator of contractility that indirectly reduces SERCA2 activity via PLN/PP1 and lowers the calcium sensitivity of myofilaments^55,57^. In contrast, CAMK2D phosphorylates RYR2, PLN, CACNA1C, and SCN5A, tending to enhance calcium handling and inotropy^55^. Importantly, there is direct genetic relevance of PRKCA and CAMK2D as demonstrated by recent DCM GWAS^34,35^.

Using an AI agent for PubMed-mining, we identified ZLN005, a small-molecule activator of peroxisome proliferator-activated receptor gamma coactivator 1-alpha (PPARGC1A,PGC-1α)^58,59^ that promotes mitochondrial biogenesis^36^. In line with prior findings in other tissues^37–39^, we demonstrated that ZLN005 reduced apoptosis also in human cardiomyocytes when exposed to injury stimuli. Mechanistically, PPARGC1A-driven mitochondrial biogenesis can bolster ATP supply and dilute ROS-damaged mitochondria. However, sustained activation of PPARGC1A increases oxygen demand and has been linked to reversible cardiomyopathy in mice^60,61^. We hypothesize that a transient pulse of mitochondrial biogenesis may support the restoration of balanced calcium handling and contractile function^62^.

Our atlas has two principal limitations. First, most snRNA-seq profiles derive from end-stage explants, which overrepresent terminal remodeling programs and insufficiently represent transient or early responses^17^. Second, genetic information was integrated from GWAS summary statistics, precluding direct variant to target gene inference from individual patients with single-cell transcriptomics. These gaps could be addressed in future work by generating matched whole-genome sequencing with snATAC-seq from the same individuals.

Beyond genetic insights, accurate clinical diagnosis, etiologic differentiation across heart-failure syndromes, and appropriate therapy selection remain critical determinants of outcomes^63^. Here, we demonstrate the feasibility of stratifying patients using multiple GWAS traits. Going forward, stratification could incorporate comorbidities and medication exposure for new therapeutic approaches.

## Data availability

Upon publication, raw sequencing files will be released on the Gene Expression Omnibus (GEO) and processed data will be made available on zenodo.

## Code availability

Code and scripts applied for the analysis of the human heart failure atlas can be found on github: https://github.com/ToreBle/Heart_Failure_Atlas

## Acknowledgments

We thank members of the Leducq Immuno-Fib Heart Failure Consortia for discussions and feedback. TB is supported by the Novo STAR postdoc grant to SH. DS was supported by the RWTH Aachen University Clinician Scientist grant. This work was funded by RWTH Aachen START (ID 692308), CRU344 and Leducq Immuno-Fib HF seed grant and Novo STAR postdoc grant to SH. RK was supported by grants from the German Research Foundation (DFG; SFBTRR219: CRU344 428857858 and CRU5011 445703531), by two grants from the European Research Council (ERC-StG 677448, ERC-CoG 101043403), a grant from the Else Kroener Fresenius Foundation (EKFS), the Dutch Kidney Foundation (DKF), TASKFORCE EP1805 and Kolff Grant no. 113351, the NWO VIDI 09150172010072 and a grant from the Leducq Foundation, and the BMBF eMed Consortium Fibromap and the BMBF Consortium CureFib.

## Contributions

TB, VD, SB, RK and SH designed the study. TB carried out the research. DS conducted validation experiments with help from CH, AA, XW and YL. JA, KH and DS did manual cell-type annotation, SM did the cell-cell communication analysis, TB and SJ worked on the GWAS association. HM sampled the human heart tissues. TB, DS, SJ, CK, MN, MB, CP, SB, VD, RK, and SH interpreted the data. TB and SH wrote the manuscript and all authors reviewed, edited and approved the manuscript.

## Methods

### Human sample acquisition and ethics

Human myocardial tissue was collected from patients undergoing heart transplantation, or implantation left ventricular assist device implantation. The use of heart tissue was approved by the local ethics committee of the Ruhr University Bochum in Bad Oeynhausen (No. 2020–640).

### Single-nuclei RNA-seq data integration and cell type annotation

For the single-cell atlas, publicly available single-nuclei RNA-seq data from different human heart failures were searched. The data sets from all studies were produced with the 10X Genomics technology by the authors. The quality of the data set was ensured by keeping nuclei with a doublet score below 2, estimated by scDblFinder^64^. Additionally, donors with at least 800 nuclei, nuclei with less than 5% of mitochondrial and ribosomal reads and a minimum of 300 features were considered. Furthermore genes were considered when they were expressed in at least 5 nuclei.

For the single-cell data integration, 5,000 highly variable genes were selected for each sample (flavor=“seurat_v3”, batch_key=“Donor”) with default settings and scVI^23^ with 2 layers and 30 latent dimensions were used.

For the main cell type annotation, different clustering resolutions of the integrated embedding were evaluated. Leiden clustering with a resolution of 0.4 was chosen for the main cell type cluster annotation and compared to the authors annotations. The main clusters were independently sub-clustered at different resolutions for the Leiden cluster and the optimal sub-clustering was chosen based on marker genes and manual inspection.

For compositional analysis, per donor aggregated cell counts for main cluster and subcluster were modeled for their relative abundances using scCODA^65^. For the distribution of different subclusters across patients, a contingency table was constructed from donor identities and subcluster annotations. Counts were z-scored across conditions to facilitate comparison of relative enrichment patterns, and hierarchical clustering was applied to group subcluster states and donor conditions with similar profiles. For the patient clustering, the counts for each patient were aggregated, integrated with harmony, clustered with Leiden and finally cell type proportion calculated from the annotations.

### Ischemic cluster evaluation

The general ischemic cell markers were obtained by differential expression analysis by the scVI differential expression module in ‘change’ mode and batch correction set to TRUE comparing the ischemic cell cluster vs all other cells of the human heart failure atlas. The differentially expressed genes were further filtered by Bayes factor>2 and non_zeros_proportion>0.05. Based on marker gene expression, Leiden resolution 0.6 was chosen for subcluster annotations of the ischemic cells.

For spatial mapping with cell2location^66^, we used the published snRNA-seq and spatial transcriptomics (Visium) datasets from Kuppe et al., 2022^16^. For the snRNA-seq data, genes and cells were filtered using a cell_count_cutoff of 5 and a cell_percentage_cutoff of 0.03, after which reference signatures of subcluster states from the heart atlas were estimated with the negative binomial regression model implemented in cell2location. These inferred reference signatures were then mapped onto spatial transcriptomics slides sampled at different sites of the human heart during acute MI. Cell2location was applied to estimate cell type abundances for each Visium spot, assuming an average of five cells per spot and using detection_alpha = 200 as a hyperparameter.

For the correction of ambient RNA, cellbender^24^ was applied to the unmapped snRNA-seq samples of acute MI^16^. Corrected count matrices were obtained with cellbender remove-background --cuda default setting (v0.3.0) and the raw_feature_bc_matrix.h5 as inputs. Subsequently, the RAW and RNA assay were loaded with Create_CellBender_Merged_Seurat from the package scCustomize (v2.0.1). Both assays were analyzed for main cell type markers, markers of as well as markers for the subclusters of the ischemic cluster using FindAllMarkers from Seurat (v5.0.1). Next, the data was subsetted for main cell types and the counterpart in the ischemic subcluster and analyzed for differentially gene expression applying FindMarkers.

Pathway analysis was performed with clusterProfiler (v4.6.2) with a minimal gene set size of 15 and max gene set size of 500. For the pathway visualizations, pathways with similar gene sets were filtered to keep the one most present across all cell types. The 10 pathways with the highest and lowest normalized enrichment score (NES) were selected, hierarchical clustered (k=8) and plotted.

Public single-cell RNA-seq data^16^ from FACS-sorted PDGFRβ CreER-tdTomato fibroblasts of mice with chronic MI by permanent ligation of the left anterior descendens artery (LAD) were re-analyzed for the gene expression of the ischemic marker.

### RNAscope experiments for co-expression validation

In situ hybridization from formalin-fixed and paraffin embedded tissue samples was performed using the RNAScope Multiplex Detection Kit v2 and 4-Plex Ancillary Kit for Multiplex Fluorescent Kit v2 (Biotechne, Cat. Nr. 323270, 323120) according to the manufacturer’s recommendations. The following probes were used for the RNAscope assay: Hs-STAB1 (Cat. Nr. 472161), Hs-SERPINE1 (Cat Nr. 555961), Hs-COL1A1-C2 (Cat. Nr. 401891-C2), Hs-PDGFRA-C3 (Cat. Nr. 604481-C3).

### Differential expression and pathway analysis

Differential expression analysis for the cell type annotation was conducted using the scVI differential expression module in ‘change’ mode and batch correction set to TRUE. The genes were further filtered by Bayes factor>2 and non_zeros_proportion>0.05. Independently, differentially expressed genes were determined from raw counts for pseudobulk analysis per sample per cluster and analyzed with DESeq2 (v1.38.3). Differentially expressed genes were considered with adjusted P<0.05 and log fold change (FC)>1 for estimating the differentially expressed genes for the different heart failures in each cell type.

Reactome pathways were obtained from the molecular signatures database (v7.5.1; https://bioconductor.org/packages/msigdb) and pathway enrichment were calculated with fgsea(minSize = 15, maxSize = 500). For the cardiomyocyte specific pathways, the differentially expressed genes were filtered for cardiomyocytes specific expression by genes with adjusted p-value<0.05 and LF>0.5 estimated from pseudobulk analysis of cardiomyocytes vs the remaining cell types.

### Cell-Cell-Communication analysis

To determine cellular interactions, ligand-receptor analysis were performed on the major cell types, considering the subclusters of ischemic cells main cluster to compare to their counterpart and excluding cell types with low abundance, namely mast cells, adipocytes, lymphatic endothelial cells and epicardium.

First, the liana consensus cell-cell-interactions (4701 ligand receptor pairs) were extended with 317 GPCR-centric ligand-receptors from the GPCR module described previously^9^. Then the *liana_wrap* function from liana (v0.1.14)^67^ was applied to each condition and Study separately. Further, the *generate_report* from CrossTalkeR (v2.0.0)^27^ function provides the p-value corrected ligand-receptor LRScores for comparison of the different heart failure diseases to control. Within each comparison the z-score and Spearman distance was calculated, followed by hierarchical clustering and the identification of top interactions of each cluster as well exclusive ones for each cluster.

In the second approach, MultiNicheNet (v1.0.3)^26^ was used to compare the ligand-receptor activity between the different heart failure diseases and controls. Prior the differential expression analysis between the conditions and cell types, genes were cut off with the following parameters: logFC_threshold = 0.50, p_val_threshold = 0.05, fraction_cutoff = 0.05. The proceeding steps, including pseudobulking, differential gene expression analysis, ligand activity prediction and prioritization were performed according to the recommendations of the vignette.

### Cardiac fibroblast *in vitro* experiments

Primary human cardiac fibroblasts (HCFs) were obtained from PromoCell (Cat. No. C-12375). Cells were cultured as recommended by the manufacturer in PromoCell Fibroblast Growth Medium 3 (Cat. No. C-23130). HCFs were seeded in 6-well plates at a density of 50,000 cells per well. After 24 hours, the medium was replaced with 0.5% FCS-containing starving medium and cultured for another 24 hours. Then, the medium was changed to serum-free medium for all wells. The cells were stimulated with either 10 ng/ml TGF-β1 (Peprotech, 100-21-100UG), 10 ng/ml IL-1β (Peprotech, 200-01B-100UG,) or hypoxic conditions (2% O2). After 24h cells were washed with PBS and RNA was isolated using the RNeasy Mini Kit (Qiagen, Cat. No. 74106) according to the manufacturer’s recommendations. NEBNext Poly(A) mRNA Magnetic Isolation Module (NEB, Cat. No. E7490) was used for mRNA enrichment and library construction was performed with the NEBNext UltraExpress RNA Library Prep Kit (NEB, Cat. No. E3330) and NEBNext Adaptor for Illumina (non-indexed), according to the manufacturer’s instructions. Bulk RNAseq samples were sequenced on an Illumina NovaSeq system targeting 25 million reads per bulkRNA library.

### Cardiomyocyte *in vitro* experiments

Human cardiac myocytes (PromoCell, C-12810) were cultivated using the Myocyte Growth Medium Kit (PromoCell, C-22170). Cells were maintained in a humidified atmosphere with 5% CO_2_ at 37°C passaged every 5-7 days using PromoCell Detach Kit (C-41210).

For total bulkRNA preparation RNA was isolated using the RNeasy Mini Kit (Qiagen, Cat. No. 74106) according to the manufacturer’s recommendations. Afterwards libraries were prepped according to the manufacturer’s recommendations using Illumina Stranded Total RNA Prep with Ribo-Zero Plus (Illumina, 20040525).

For the experiments, 60 000 cells were seeded in 8-well plates (Ibidi, Cat. No. 80827) or 6-well plates (Greiner, 657160) in Myocyte Growth Medium supplemented with 1% FCS (PromoCell, C-22170). The next day, cells were washed with PBS and treated with their corresponding experimental condition for 48 h. Treatments were prepared in serum free medium containing either 10 µM ZLN005 (MedChem Express, HY-17538-100mg, dissolved in DMSO (AppliChem, A3672)) or the correlating amount of DMSO as vehicle control. For stimulation, the cells were exposed to either 10 ng/ml recombinant human TGF-β (PeproTech, 100-21), 10 ng/ml IL-1β (PeproTech, 200-01B-100UG) or 1 µM doxorubicin (Sigma-Aldrich, D5220-1ML). Hypoxic conditions were induced by incubating the cells in a hypoxia chamber maintained at 2% O_2_.

### Bulk-RNA seq alignment, DEG, pathway analysis and deconvolution

For the bulkRNA sequencing, STAR (v2.7.11b) was used for alignment to the human genome (GRCh38 v98), followed by removal of duplicated reads with Picard, estimation of gene-level counts by featureCounts, differential expression analysis by DESeq2 and pathway enrichment with gene set enrichment analysis and reactome pathways using clusterProfiler.

To link each fibroblast stimulations to fibroblast subclusters from the heart atlas, a reference profile for each subcluster was estimated from the single-cell data and Bisque^28^ were applied to estimate subcluster contributions per bulk RNA-seq sample.

### Integration of genetics from GWAS summary statistics

The NHGRI-EBI GWAS Catalog was queried for studies related to heart failure, vascular diseases, relevant comorbidities, and associated clinical measurements. GWAS with the largest discovery sample size, broad ancestral diversity and available summary statistics for hg38 were prioritized. Furthermore, two recent studies for HCM^68^ and DCM^34^ were added.

For the integration, scPagwas (v1.3.0)^30^ which links single cells and cell types to complex traits by modeling pathways while preserving per-cell resolution. For non-failing donors, HCM and DCM patients the scPagwas analysis was performed on a patient level by firstly mapping SNPs from given GWAS summary statistics to nearby genes and then grouping them into pathway blocks. For each pathway of the provided reduced reactome pathway set, scPagwas fits a polygenic regression that relates LD-aware SNP effect sizes to pathway-level activity derived from single-cell expression, producing per-cell, per-pathway genetic contribution estimates defined as the scPagwasPaHeritability assay. From this Seurat assay the mean pathway proportions and cell type specificity were estimated. Then summing these across pathways yields a genetic pathway activation score (gPAS) for each cell. To highlight genes driving the per-cell genetic signal, a Pearson correlation coefficient (PCC) for every gene between its expression and the cell’s gPAS was calculated. Finally, a Trait Relevance Score (TRS) is calculated per cell by scoring modules of the top PCC genes using Seurat’s AddModuleScore. For the patient stratification, the mean TRS for each cell type across patients with DCM and individuals with non-failing hearts was calculated from selected GWAS summary statistics. Patient and cell type specific TRS were subsetted for cardiomyocytes, clustered and plotted with selected meta data using ComplexHeatmap (v2.14.0).

GTEX v8 (ENCODE v26) was leveraged for the expression Quantitative Trait Locus (eQTL) analysis. The GWAS summary statistics for DCM^34^ were integrated by SNP IDs with eQTL data from selected human adult tissue. The colocalization was assessed for candidate genes using eQTpLot and based on the matching coordinates for hg38 ATAC-seq^10^ of selected human cardiac cell types were visualized.

### AI agent for pathway based drug target discovery

An AI agent was set up with chatGPT4.1 and Claude Sonnet 4. The agent reads a JSON file with a subset of relevant pathways obtained from the heart atlas and processes a PubMed search query based on a research context and the gene list pre-defined relevant for the pathway. The query was submitted to the currently available PubMed MCP server (https://github.com/cyanheads/pubmed-mcp-server), requesting a ranking by publication date for results in between the years of 2015 and 2025 with limiting fetching of 882 maxResults and 42 fetchBriefSummaries, addressing the limits of PubMed article fetching criteria. Furthermore, the agent was instructed to check the presence of identified chemical compounds from the PubMed search with the search_compound tool, querying a currently available PubChem MCP server (https://github.com/Augmented-Nature/PubChem-MCP-Server). The outputs of the agent were saved to a .json file and subsequently plotted with ggplot2 and ggwordcloud.

### Tunel staining

Apoptotic cardiac myocytes were detected using the In Situ Cell Death Detection Kit, TMR red (Roche, Cat. No. 12156792910) according to the manufacturer’s instruction (version 13) for adherent cells on 8-well glass bottom slides (Ibidi, Cat. No. 80827). Cells were fixed with 4% paraformaldehyde (Roth, P087.3) for one hour at room temperature, washed with PBS (Bio&Sell, B.S.L 1825) and permeabilized with 0.1% Triton X-100 (Sigma-Aldrich, T8787) in PBS for two minutes on ice. After washing, samples were incubated with the TUNEL reaction mix for 60 minutes at 37°C in a humidified dark chamber, rinsed with PBS and counterstained with DAPI (1:1000 in RNase free water (Qiagen, 1017979) for 10 minutes at 37°C in the dark. Negative controls were prepared by replacing the reaction mixture with label solution, while positive controls were generated by treating cells overnight with 0.05mM staurosporine (Selleck Chemicals, S1421 in Myocyte Growth Medium (PromoCell, C-22170)).

Fluorescence images were acquired on a ZEISS LSM 980 confocal microscope operated with ZEN software, using a 10x objective, a 405nm laser for DAPI and 561 nm laser for the Cy3/TMR red. Images were processed and analysed in QuPath (v0.6.0). Cell detection was based on DAPI stained nuclei; apoptotic cells were identified using QuPath’s Positive cell detection tool. For every replicate five fields of view were analysed. The results were given as absolute counts per image, and the percentage of positive cells was calculated for each condition.

### RNA isolation, cDNA synthesis and quantitative RT-PCR

Cells were washed with PBS followed by RNA extraction according to the manufacturer’s protocol using the RNeasy Mini Kit (Qiagen, Cat. No. 74106). 200 ng total RNA was reverse transcribed with High-Capacity cDNA Reverse Transcription Kit (Thermo Fisher Scientific, Cat. No. 4368813) and qRT-PCR was carried out using iTAQ SYBR Green Supermix (Bio-Rad, Cat. No.. 1725125) and primers for *bACTIN* (CACCATTGGCAATGAGCGGTTC, AGGTCTTTGCGGATGTCCACGT), *NPPB* (TCTGGCTGCTTTGGGAGGAAGA, CCTTGTGGAATCAGAAGCAGGTG) and *ANKRD1* (CGACTCCTGATTATGTATGGCGC, GCTTTGGTTCCATTCTGCCAGTG). Data were analyzed using the 2-CT method.

