## Supplementary Figures for "Integrative single-cell and genetic profiling of human heart failure identifies targets for cardiomyocyte restoration"

### Supplementary Figure 1

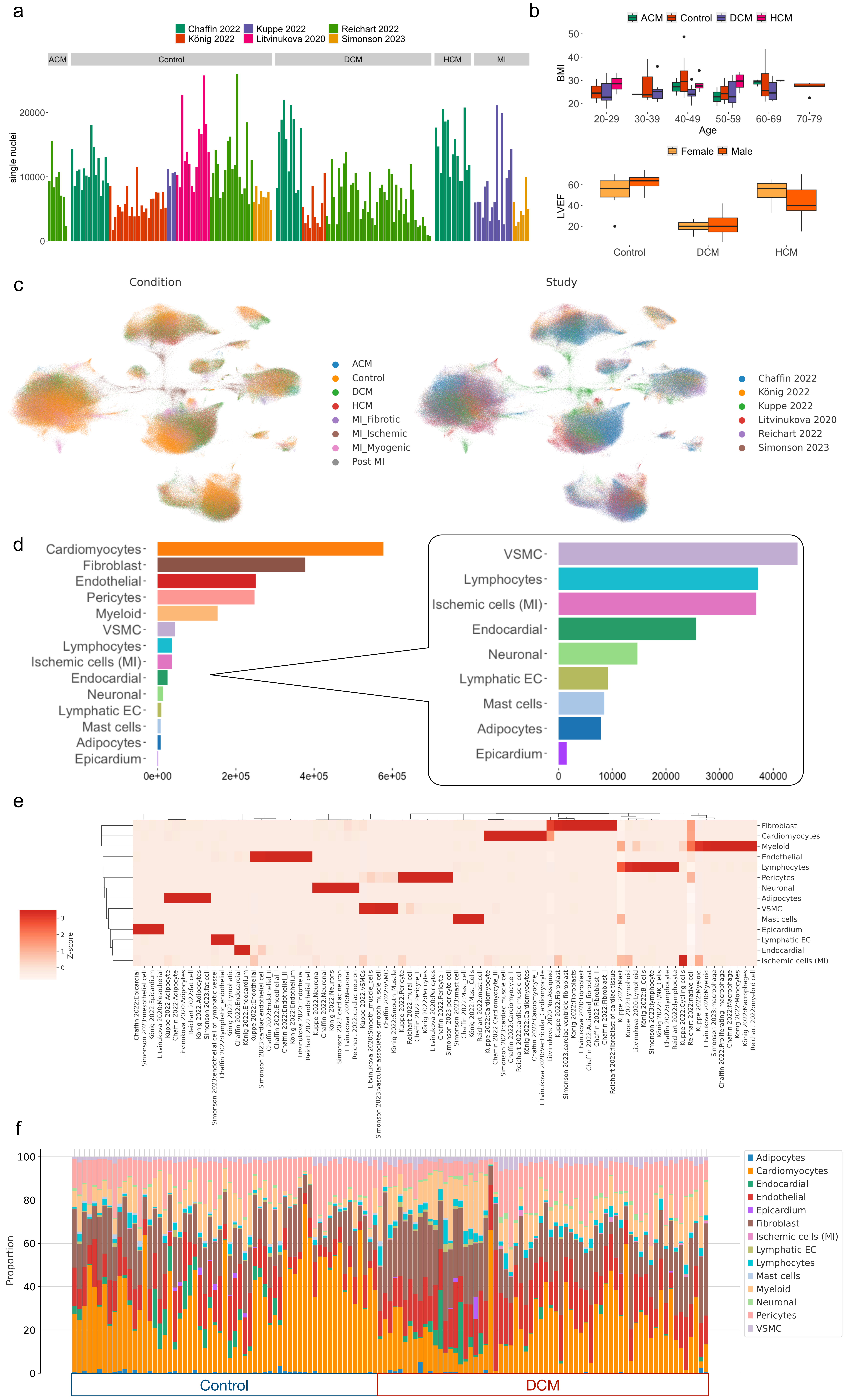

### Supplementary Figure 2

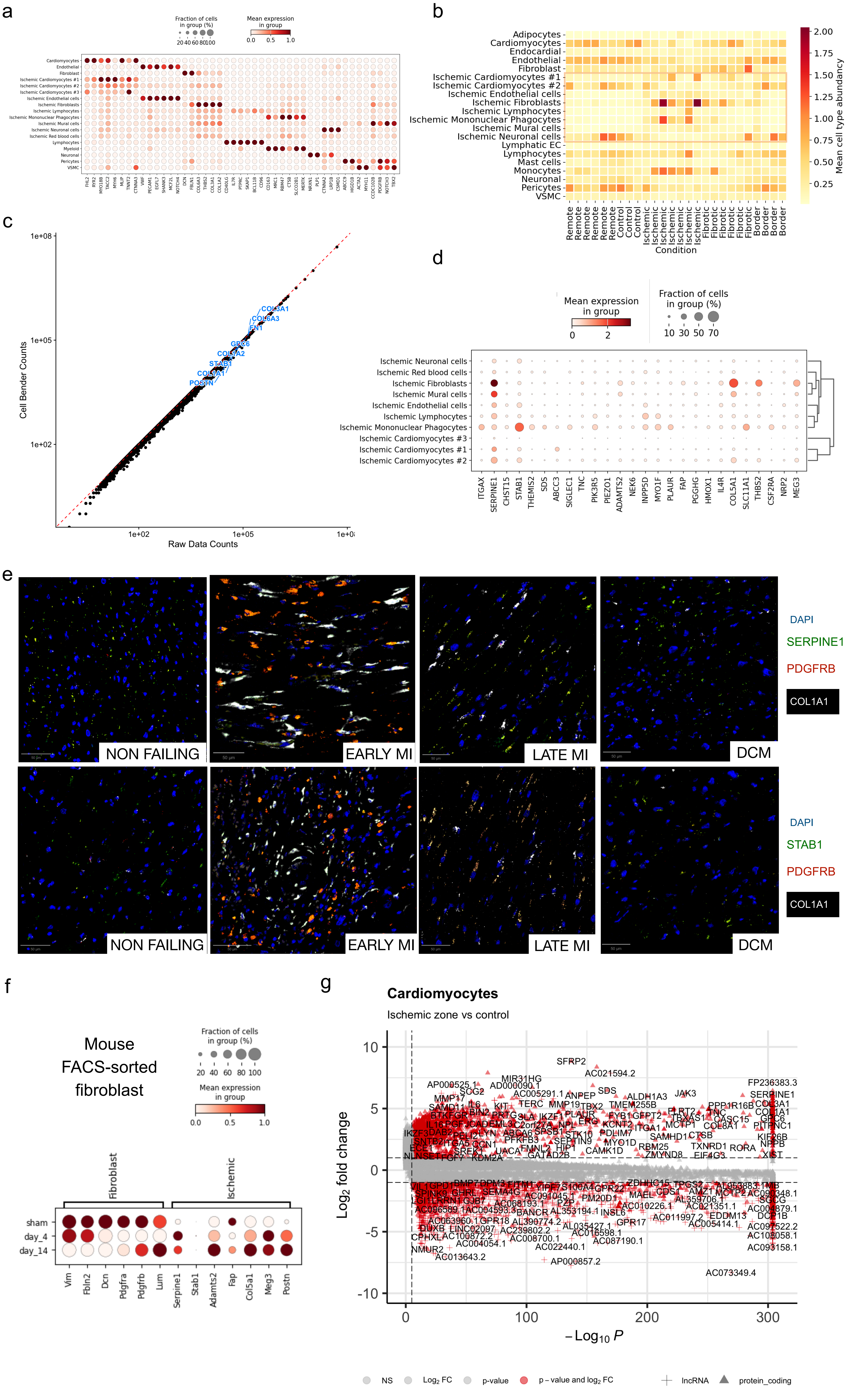

### Supplementary Figure 3

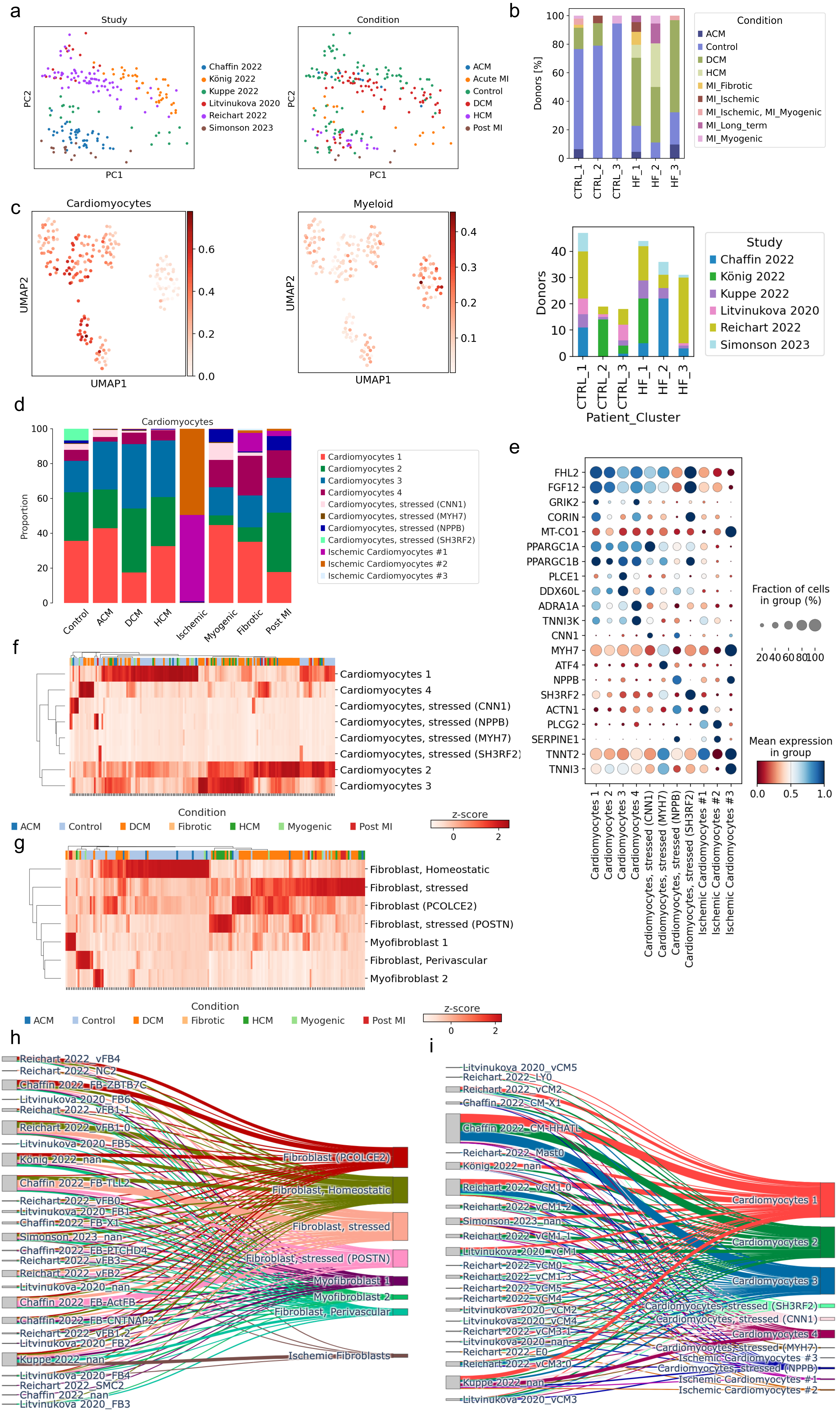

### Supplementary Figure 4

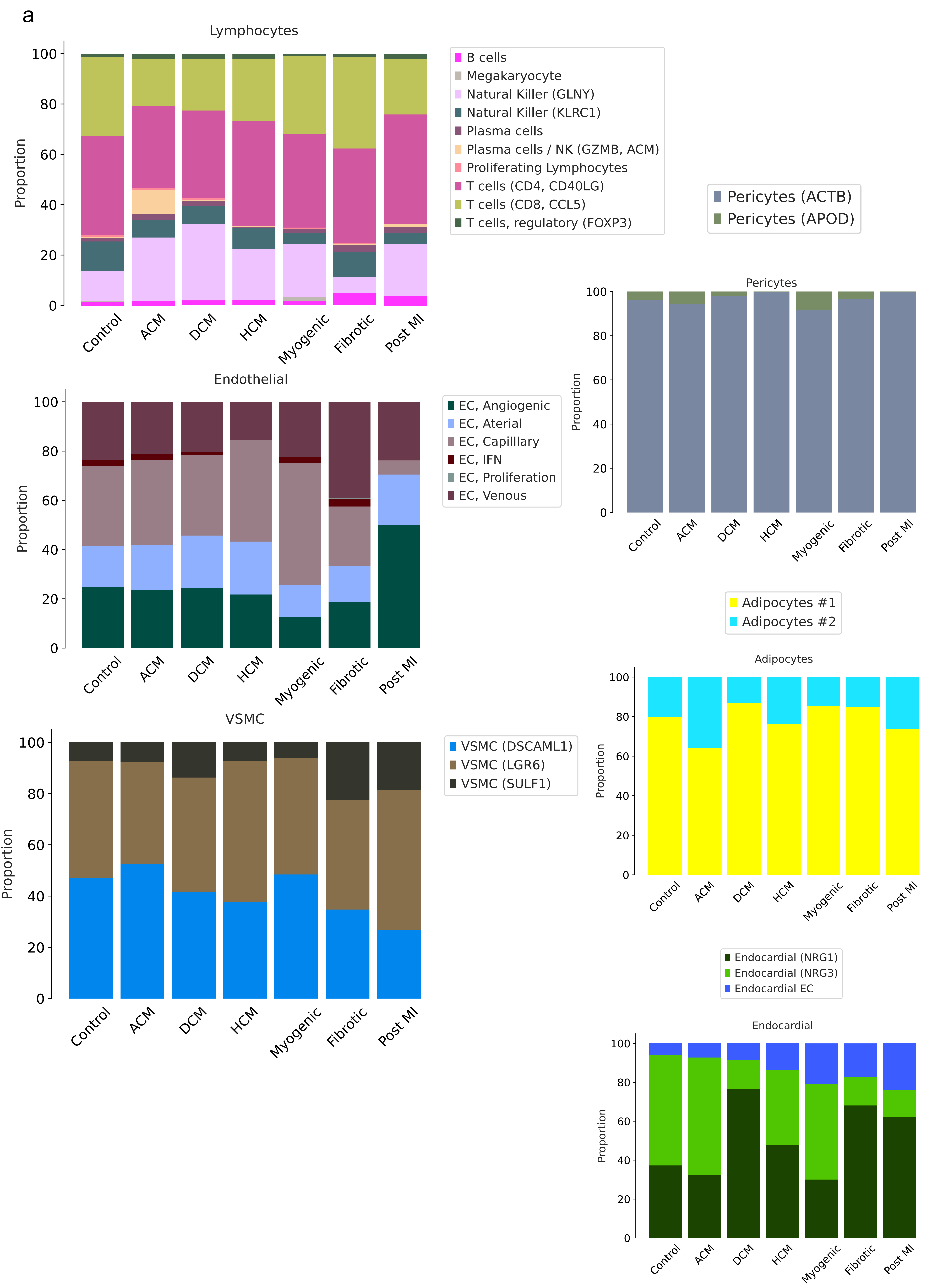

### Supplementary Figure 5

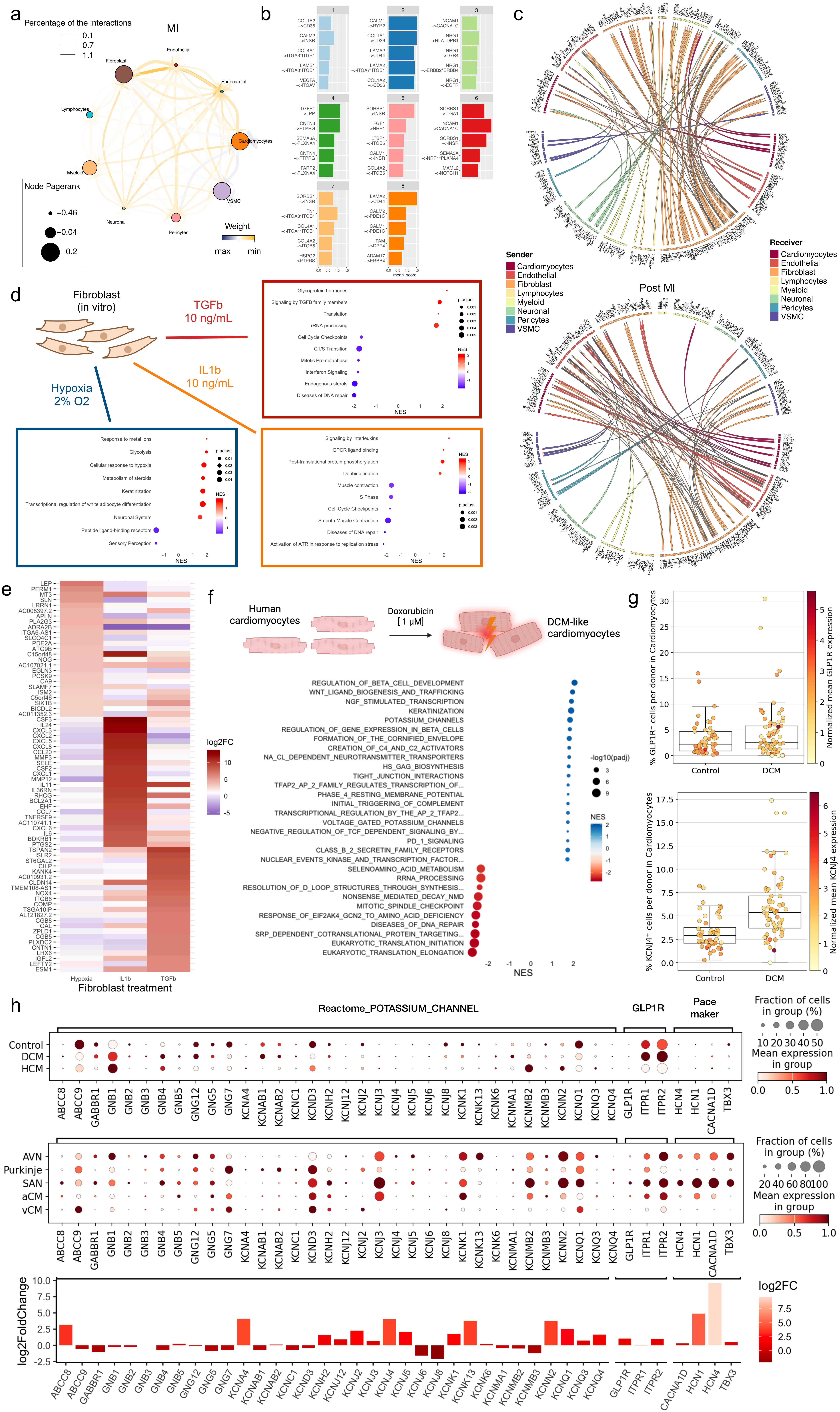

### Supplementary Figure 6

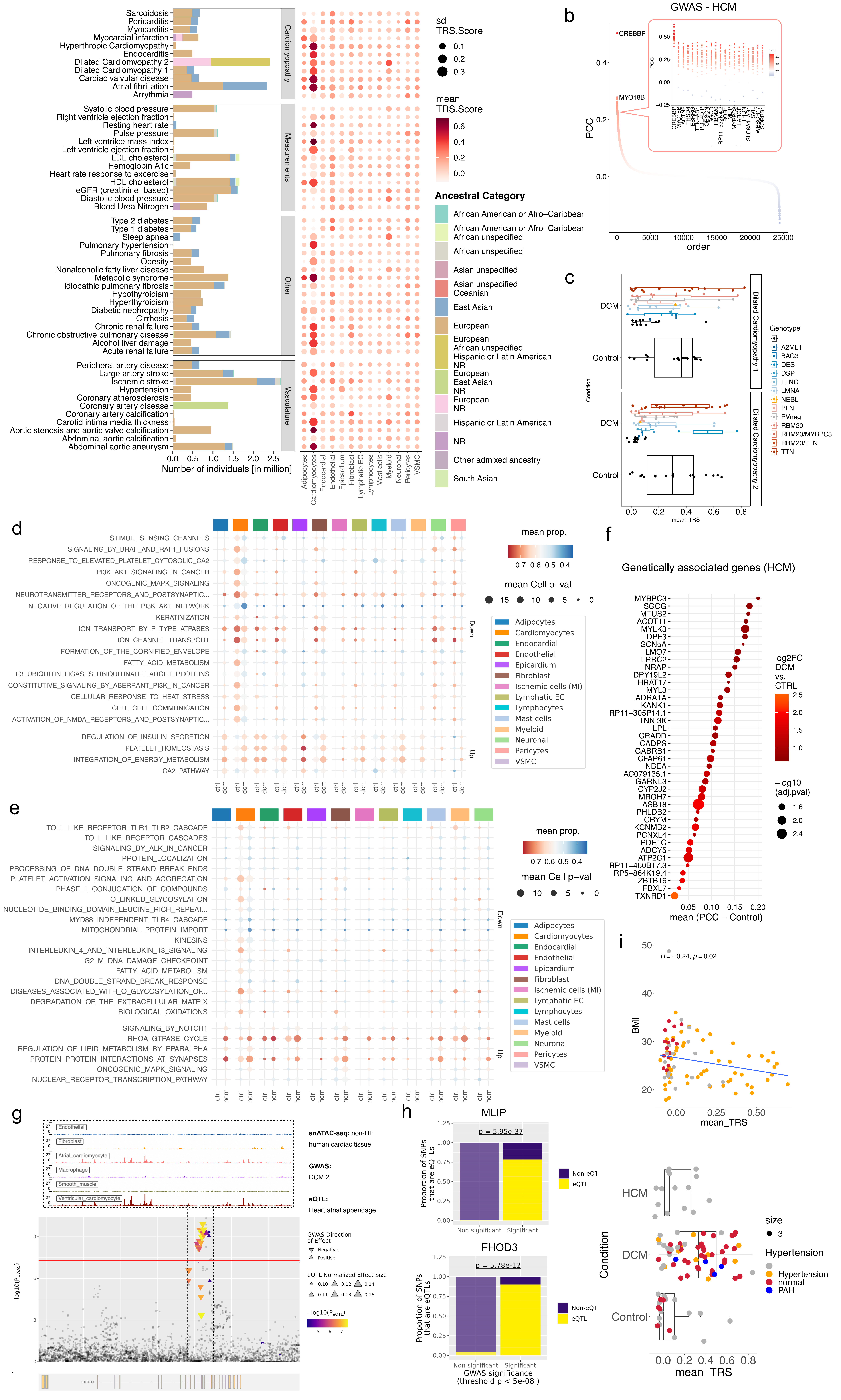

### Supplementary Figure 7

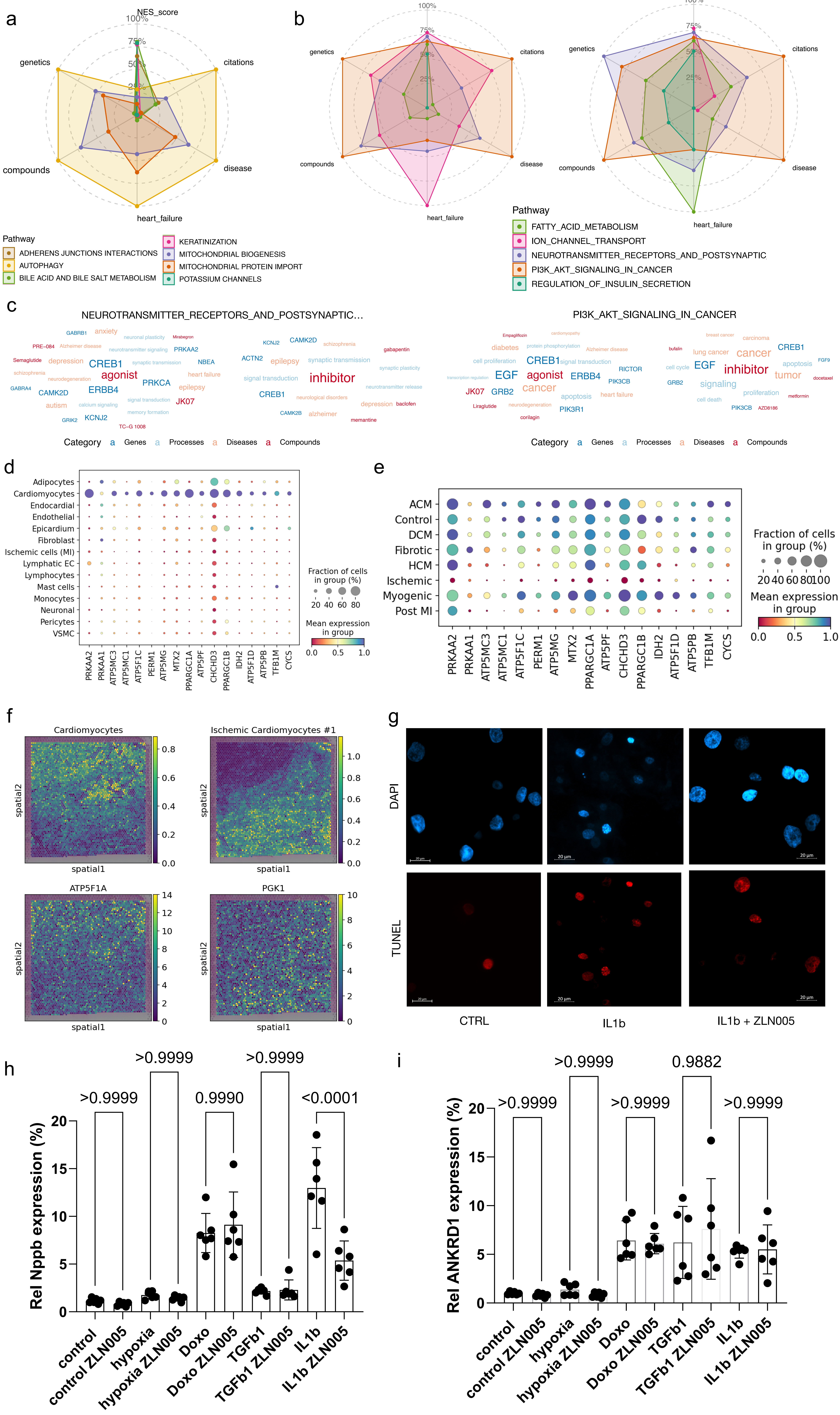

#### Supplementary Figures

##### Figure 1: Integration of the human heart failure atlas

a - Overview of the numbers of nuclei per donor, grouped by the heart failure condition and colored by the study the data were published in. ACM: arrhythmogenic cardiomyopathy; DCM; dilated cardiomyopathy; HCM: Hypertrophic cardiomyopathy; MI: myocardial infarction.

b - Boxplot showing the body-mass-index (BMI) by age groups and different cardiomyopathies.

c - UMAP of the integrated human heart failure atlas colored by the heart failure condition (left) and the original study of the data (right).

d - Cross table of the major cell type annotation of the integrated human heart failure atlas (y-axis) and the original annotations of each study. The color shows the z-normalized proportion for each author annotation.

e - Comparison of z-scores for the major cell type annotations obtained from the individual studies that were integrated into the human heart failure atlas and the unified annotation in our integrated atlas.

f - scCODA defined cellular composition proportions of the major cell types by the heart donor patients (control) and DCM patients.

##### Figure 2: Gene signature of the ischemic cell cluster

a - Differential expression within the ischemic clusters showing the indicated markers for the different cell types. For comparison, the major cluster marker from Fig. 1e for the most similar cell type are shown as well.

b - Spatial cell type abundance of the mapped cell stated in different Visium slides from control donors and heart tissue sites from patients with acute myocardial infarction. The 10X Genomics Visium slides were analyzed from Kuppe *et al.*, 2022.

c - Scatterplot of the gene expression counts defined by the raw counts and ambient RNA-adjusted counts using CellBender.

d - Marker gene expression of the ischemic cell cluster within the subtypes of the ischemic cluster (MI).

e - Representative images from RNAscope experiments from *SERPINE1* (top) and *STAB1* (bottom) for non-failing controls, early and late MI as well as DCM. Each experiment includes DAPI stainings and RNAscope for *COL1A1* and *PDGFRB*.

f - Expression of fibroblast and ischemic marker in single-cell RNA-seq from FACS-sorted PDGFRβ CreER-tdTomato MI mice (n=4154 (sham); n = 13263 (day_4); n = 21554 (day_14).

g - Volcano plot of the differential expression analysis of ischemic cardiomyocytes vs cardiomyocytes from control samples.

**Figure 3: SnRNA based sample stratification and cell type subclustering**

a - PCA of the pseudobulk aggregated patients colored by study and condition.

b - Composition of the patient clusters by condition (top) and study (bottom).

c - UMAP of the patient clustering colored by the percentage of the cardiomyocytes (left) and myeloid (right) cluster in each patient snRNA-seq sample.

d - *scCODA*-based compositional analysis of cardiomyocyte cell subcluster across conditions, including non-failing controls, cardiomyopathies, and myocardial infarction at different stages.

e - Gene expression of selected markers for each cardiomyocyte subcluster.

f - z-score of individuals for the cardiomyocyte subcluster of the human heart failure atlas

g - z-score of individuals for the fibroblast subcluster of the human heart failure atlas.

h - Sankey plot of the fibroblast subclusters showing the annotations in individual studies on the left and the unified fibroblast subcluster annotation of the human heart failure atlas on the right.

i - Sankey plot of the cardiomyocyte subclusters showing the annotations in individual studies on the left and the unified cardiomyocyte subcluster annotation of the human heart failure atlas on the right.

**Figure 4: Cardiac cell type subclustering and differentially gene expression**

a - *scCODA*-based compositional analysis of lymphocyte, endothelial cell,endocardial, pericytes, vascular smooth muscle cells (VSMC) and adipocytes subcluster across conditions, including non-failing controls, cardiomyopathies, and myocardial infarction at different stages.

**Figure 5: Cell-Cell-Comminaction analysis and signal influence on different fibroblast subcluster states**

a - CrosstalkR results for the cell-cell-communication in the different heart failure conditions. Shown are the weighted interactions for MI. For the analysis the consensus interactome of liana was extended by the GPCR module.

b - Top ligand and receptors for each interactome cluster by LRScore.

c - MultiNicheNet prioritized ligand activities for MultiNicheNet prioritized ligand activities for DCM and post myocardial infarction in the major cardiac cell types.

d - *in vitro* fibroblast differentiation with different treatments followed by bulk RNA-sequencing. The normalized pathway enrichment score (NES) was calculated for each condition vs. control from 6 biological replicates.

e - *in vitro* fibroblast differentiation with different treatments followed by bulk RNA-sequencing. Shown are the top differential expressed genes from DESeq2 for each condition vs. control from 6 biological replicates.

f - Overview of the bulkRNA-seq experiment of *in vitro* cultured human cardiomyocytes and their treatment with 1 µM doxorubicin to a DCM-like state. Following differential expression analysis treatment and control (n=3), fsgsea was applied for reactome pathway analysis which is colored by the normalized enrichment score (NES) and the size of the dots represents the -log10 of the adjusted p-values.

g - Gene expression of *GLP1R* (top) and *KCNJ4* (bottom) within cardiomyocytes of individuals. Shown are the percentage of cardiomyocytes expressing the gene and the mean expression of the gene within those cells.

h - Gene expression of genes from the reactome pathway POTASSIUM_CHANNELS in cardiomyocytes. Shown are genes expressed in minimum of 3% of the cardiomyocytes of the heart atlas as well as non-overlapping selected genes from the GLUCAGON_LIKE_PEPTIDE_1_GLP1_REGULATES_INSULIN_SECRETION and known pacemaker genes. The top shows the expression of the genes in the different conditions of cardiomyopathies, the middle within the cardiac niches using the dataset from Kanemaru et al., 2023. The bottom shows the log2FoldChange from the *in vitro* treatment of human cardiomyocytes with 1 µM doxorubicin.

#### Figure 6: Linkage of single-cell expression and genetics

a - Overview of the analyzed GWAS traits showing the number of individuals and the ancestry for each trait grouped by cardiomyopathies, vasculature and other diseases traits as well as traits to related clinical measurements on the left and the trait-relevant scores for all analyzed GWAS traits on the right. The TRS score was calculated per patient. Shown are the mean value per cell type cluster from the patient of the non-HF control as well as dilative and hypertrophic cardiomyopathy. The size denotes the standard deviation (sd) of the mean TRS score.

b - HCM-relevant genes ranked by the Pearson correlation coefficient (PCC) as mean from patients with non-failing hearts. The top ranked genes are highlighted in the box, where each dot denotes the ranking and PCC from individual samples.

c - Comparison of the trait-relevant score (TRS) for the GWAS DCM 1 and DCM 2 in cardiomyocytes for DCM samples highlighted by the genotype (black/empty refers to patients that were not genotyped) and samples from the non-HF control group.

d - Trait-relevant pathways for DCM and cardiomyocytes showing the proportion of cells with the indicated pathway for each cell type and the condition. The color indicates the mean proportion from the individual patients (pathway-level coefficient $\beta > 0$) for each cell type.

e - Trait-relevant pathways for HCM and cardiomyocytes showing the proportion of cells with the indicated pathway for each cell type and the condition. The color indicates the mean proportion from the individual patients (pathway-level coefficient $\beta > 0$) for each cell type.

f - Genetically enriched genes and their changes in PCC for the GWAS of HCM.

g - Manhattan plot for -/+ 200kb of the FHOD3 gene for the GWAS for DCM 2. Overlapping SNPs with the expression quantitative trait locus (eQTL) from the heart atrial appendage are highlighted by the effect size. For the dashed region with significant SNP, the single-nuclei ATAC-seq data from Hocker et al. is shown for the indicated cell types.

h - Enrichment of eQTLs among GWAS significant SNPs. Compared are proportions of overlapping SNPs from the eQTLs and GWAS SNPs from DCM 2 of *MLIP* and *FHOD3*.

i - Pearson correlation analysis of the average trait-relevant score (TRS) for the obesity trait in cardiomyocytes versus the body-mass-index (BMI) of the patients with DCM and non-HF and the comparison of the TRS for the GWAS of hypertension in cardiomyocytes for DCM samples and control highlighted by the meta data related to hypertension (gray/empty refers to patients without clinical meta data for hypertension).

**Figure 7: Drug target discovery and experimental validation for cardiomyocyte rescue**

a - Radar plot showing the analysis of the AI agent output from the pubmed search for cardiomyocyte-specific pathways in heart failure in the research context of inhibitors. The total occurrences of disease, heart failure, chemical compounds and genetic related studies as well as the overall number of studies (citations) are shown min-max normalized, while the NES was scaled from -3 (equal to 0%) to +3 (equal to 100 %).

b - Radar plot showing the analysis of the AI agent output from the pubmed search for DCM GWAS associated pathways and their mean proportion estimated by scPagwas with the research context for agonists (left) and inhibitors (right). The total occurrences of disease, heart failure, chemical compounds and genetic related studies as well as the overall number of studies (citations) are shown min-max normalized.

c - Word frequency plots derived from the AI agent PubMed search of the genetically associated pathways for neurotransmitter receptors and postsynaptic signal transmission and PI3K AKT signaling in cancer colored by the different categories.

d - Gene expression of genes associated with the reactome pathways of mitochondrial biogenesis within the major cell types of the human heart atlas.

e - Gene expression of genes associated with the reactome pathways of mitochondrial biogenesis within cardiomyocytes by the indicated conditions.

f - Metabolic changes at the MI border represented by the expression of ATP synthase F1 subunit alpha (*ATP5F1A)* for oxidative phosphorylation in cardiomyocytes and phosphoglycerate kinase (*PGK1*), an inhibitor of the oxidative phosphorylation, for glycolysis in ischemic cardiomyocytes.

g - Representative DAPI (top) and TUNEL staining (bottom) of untreated cardiomyocytes (control), cardiomyocytes treated with IL-1β and IL-1β and the PGC-1α agonist ZLN005.

h - RT-qPCR results showing the normalized expression of *NPPB* for the cardiomyocyte treatment with IL-1β and doxorubicin and the addition of ZLN005. On top are displayed the p-values for the treatment and the recovery with ZLN005 from a one-way ANOVA for multiple comparisons.

i - RT-qPCR results showing the normalized expression of *ANKRD1* for the cardiomyocyte treatment with IL-1β and doxorubicin and the addition of ZLN005. On top are displayed the p-values for the treatment and the recovery with ZLN005 from a one-way ANOVA for multiple comparisons.
